# Predicting mortality, duration of treatment, pulmonary embolism and required ceiling of ventilatory support for COVID-19 inpatients: A Machine-Learning Approach

**DOI:** 10.1101/2021.02.15.21251752

**Authors:** Abhinav Vepa, Amer Saleem, Kambiz Rakhshan, Amr Omar, Diana Dharmaraj, Junaid Sami, Shital Parekh, Mohamed Ibrahim, Mohammed Raza, Poonam Kapila, Prithwiraj Chakrabarti, Tabassom Sedighi, Omid Chatrabgoun, Alireza Daneshkhah

## Abstract

**Introduction:** Within the UK, COVID-19 has contributed towards over 103,000 deaths. Multiple risk factors for COVID-19 have been identified including various demographics, co-morbidities, biochemical parameters, and physical assessment findings. However, using this vast data to improve clinical care has proven challenging.

**Aims:** to develop a reliable, multivariable predictive model for COVID-19 in-patient outcomes, to aid risk-stratification and earlier clinical decision-making.

**Methods:** Anonymized data regarding 44 independent predictor variables of 355 adults diagnosed with COVID-19, at a UK hospital, was manually extracted from electronic patient records for retrospective, case-controlled analysis. Primary outcomes included inpatient mortality, level of ventilatory support and oxygen therapy required, and duration of inpatient treatment. Secondary pulmonary embolism was the only secondary outcome. After balancing data, key variables were feature selected for each outcome using random forests. Predictive models were created using Bayesian Networks, and cross-validated.

**Results:** Our multivariable models were able to predict, using feature selected risk factors, the probability of inpatient mortality (F1 score 83.7%, PPV 82%, NPV 67.9%); level of ventilatory support required (F1 score varies from 55.8% “High-flow Oxygen level” to 71.5% “ITU-Admission level”); duration of inpatient treatment (varies from 46.7% for “≥ 2 *days but* < 3 *days*” to 69.8% “≤ 1 *day*”); and risk of pulmonary embolism sequelae (F1 score 85.8%, PPV of 83.7%, and NPV of 80.9%).

**Conclusion:** Overall, our findings demonstrate reliable, multivariable predictive models for 4 outcomes, that utilize readily available clinical information for COVID-19 adult inpatients. Further research is required to externally validate our models and demonstrate their utility as clinical decision-making tools.

**Highlights:**

- Using COVID-19 risk-factor data to assist clinical decision making is a challenge
- Anonymous data from 355 COVID-19 inpatients was collected & balanced
- Key independent variables were feature selected for 4 different outcomes
- Accurate, multi-variable predictive models were computed, using Bayesian Networks
- Future research should externally validate our models & demonstrate clinical utility

## 1. Introduction

On Thursday the 5^th^ of March 2020, within the UK, COVID-19 claimed the life of its first victim and has since contributed towards over 103,000 deaths [1,2]. The Office of National Statistics (ONS) has since issued statements, based on population data, in conjunction with the National Health Service (NHS), indicating an increased risk of mortality from COVID-19 amongst poorer socioeconomic groups, Black and Minority Ethnics (BAME), males and the elderly [3–6]. In addition to demographics, various biochemical parameters and co-morbidities, such as obesity, diabetes, hypertension, chronic obstructive pulmonary disease (COPD) and malignancy, have been identified as risk factors for poor COVID-19 outcomes [7–10]. However, using this vast data to improve clinical care has proven challenging. One particular challenge that remains is relatively quantifying the impact of various prognostic indicators on COVID-19 outcomes, especially whilst in the presence of combinations of other variables, in order to assist clinical decision making and risk stratification.

Due to the limited nature of healthcare resources, such as hospital beds and ventilators, clinicians are often faced with difficult decisions where they must ration resources between patients, often having ethical implications [11]. Currently, clinicians are allocating healthcare resources to COVID-19 patients semi-quantitatively, and often as a response to clinical deterioration. Various risk stratification models have been described in the literature such as the 4C tool [12], but are currently not being used clinically due to criticism in recent systematic reviews [13,14]. Some of the key problems with existing risk-stratification tools are unclear methodologies, the exclusion of patients diagnosed with COVID-19 using CT imaging but with negative Real Time-Polymerase Chain Reaction (RT-PCR) nasopharyngeal swabs, small sample sizes, many patients not reaching a study outcome, automated data extraction relying on clinical coding and many studies only exploring inpatient mortality as a primary outcome. In addition, many predictive models have been developed using patient data from other parts of the world, which may not be generalizable to the UK population due to patient factors, hospital factors and virus factors. Finally, only a small selection of risk-stratification tools analysed a wide host of independent variables including vital observations, biochemical markers, demographics, and co-morbidities.

The aim of this study was to develop a reliable, multivariable, predictive model, which utilizes readily available clinical data, to serve as a quantitative tool to aid risk-stratification, and earlier clinical decision-making for adult, hospitalized COVID-19 patients.

## 2. Methods

### 2.1 Study Design and Setting

This retrospective, case-controlled study was conducted at Milton Keynes University Hospital (MKUH), which is a medium sized, 550 bed, and district general hospital in the United Kingdom. Data was collected during the routine clinical care of patients for auditing purposes, and upon receiving Health Research Authority (HRA) approval, the anonymous data was then also used for research purposes. The study aimed to follow the Transparent Reporting of a multivariable prediction model for Individual Prediction of Diagnosis (TRIPOD) checklist [15] and was conducted according to a pre-defined study protocol.

### 2.2 Study Population

Adult patients diagnosed with positive RT-PCR nasopharyngeal swabs or Computed Tomography (CT) scans with changes suggestive of COVID-19 [16], between 01/03/2020 and 22/04/2020 at MKUH, were included in this study. 69 patients were excluded which is shown below in Table 1, to produce a final n number of 355. The sample size was determined by using the maximum number of COVID-19 patients diagnosed during the study period. The study end date was the date of initiating independent predictor variable data collection (22/4/2020) whereas the study start date was the date of the first COVID-19 patient diagnosis (1/3/2020). Patient characteristic information is shown in Supplementary Table 1.

**Table 1:** Patient selection process

| **Sample Population** |  |
| --- | --- |
| Patients diagnosed with COVID-19 between 01/03/2020 and 22/04/2020, at Milton Keynes University Hospital (n=424) |  |
| **Inclusion Criteria** | **Exclusion Criteria** |
| 1. Patients diagnosed with at least 1 positive RT-PCR Nasopharyngeal swab<br>2. Patients diagnosed with CT scan changes consistent with COVID-19 based on BSTI Guidance [17]<br>3. Age 18 years and above | 1. Patients diagnosed in the Outpatient setting<br>2. Staff Members who were diagnosed via Occupational Health, and who did not receive an official medical assessment |
| Final Study Participant Number (n = 355) |  |

**Table:1.** List and definitions of independent predictor variables

| Independent Predictor Variables (p=44) | Definition |
| --- | --- |
| Age | The number of completed years since self-reported date of birth (years) |
| Gender | Self-reported gender by study participant (male or female) |
| Ethnicity | Self-reported ethnicity by study participant (Caucasian or BAME). 'Mixed' ethnicities and 'other' ethnicities were classified as BAME. |
| Oxygen Saturations | The lowest oxygen saturation reading obtained by nursing staff on the day of COVID-19 clinical presentation whilst an inpatient (%) |
| Respiratory Rate | The highest respiratory rate reading obtained by nursing staff on the day of COVID-19 clinical presentation whilst an inpatient (breaths per minute) |
| Temperature | The highest temperature reading obtained by nursing staff on the day of COVID-19 clinical presentation whilst an inpatient (°Celsius to 1 decimal point) |
| Obesity | A body mass index more than 30, or a clinical diagnosis of obesity documented within the patients past medical history (Yes or No) |
| Previous Venous Thromboembolism | A confirmed clinical diagnosis of any venous thromboembolism documented within the patients past medical history or a patient who is prescribed a treatment dose of anticoagulation (Yes or No) |
| Chronic Obstructive Pulmonary Disease | A confirmed clinical diagnosis of emphysema, bronchitis, or chronic obstructive pulmonary disease documented within the patients past medical history, or, a study participant with smoking history who is prescribed a long-term bronchodilator or corticosteroid inhaler (Yes or No) |
| Bronchiectasis | A confirmed clinical diagnosis of bronchiectasis documented within the patients past medical history (Yes or No) |
| Asthma | A confirmed clinical diagnosis of asthma documented within the patients past medical history (Yes or No) |
| Interstitial Lung Disease | A confirmed clinical diagnosis of interstitial lung disease or pulmonary fibrosis documented within the patients past medical history (Yes or No) |
| Lung Cancer | A confirmed clinical diagnosis of lung cancer documented within the patients past medical history (Yes or No) |
| Neuromuscular Disease | Any neurological or neuromuscular disease confirmed by clinical diagnosis documented within the patients past medical history such as strokes, epilepsy, and Parkinson disease (Yes or No) |
| Diabetes Mellitus | A confirmed clinical diagnosis of diabetes mellitus documented within the patients past medical history or a study participant who is prescribed long-term oral hypoglycaemic drugs or insulin (Yes or No) |
| Hypertension | A confirmed clinical diagnosis of hypertension documented within the patients past medical history or a study participant who is prescribed long-term antihypertensive drugs (Yes or No) |
| Ischaemic Heart Disease | A confirmed clinical diagnosis of ischaemic heart disease documented within the patients past medical history or a study participant who is prescribed a long-term antiplatelet drug (Yes or No) |
| Chronic Kidney Disease | A confirmed clinical diagnosis of chronic kidney disease documented within the patients past medical history or a baseline estimated glomerular filtration rate of < 60 (Yes or No) |
| Non-steroidal anti-inflammatory drugs | Repeat prescription, within the last 3 months, of oral non-steroidal anti-inflammatory drugs documented within the patient's drug history (Yes or No) |
| Anticoagulant | Repeat prescription, within the last 3 months, of anticoagulation documented within the patient's drug history (Yes or No) |
| Antibiotics | Repeat prescription, within the last 3 months, of antibiotics documented within the patient's drug history (Yes or No) |
| Immunosuppressants | Repeat prescription, within the last 3 months, of immunosuppressants, excluding corticosteroids and previous completed chemotherapy, documented within the patient's drug history (Yes or No) |
| Corticosteroids | Repeat prescription, within the last 3 months, of oral corticosteroids documented within the patient's drug history (Yes or No) |
| Angiotensin Converting Enzyme Inhibitors | Repeat prescription, within the last 3 months, of angiotensin converting enzyme inhibitors documented within the patient's drug history (Yes or No) |

| **Independent Predictor Variables (p=44)** | **Definition** |
| --- | --- |
| Angiotensin Receptor Blockers | Repeat prescription, within the last 3 months, of angiotensin receptor blockers documented within the patient's drug history (Yes or No) |
| CT imaging severity of COVID-19 related changes | Extent of air-space consolidation or ground-glass opacification in lung tissue on CT scans, where available, performed during inpatient stay (unilateral or bilateral) |
| COVID-19 related Chest X-Ray changes | Shadowing, consolidation, or haziness attributed to COVID-19 by a consultant radiologist on a chest X-Ray, where available, performed during inpatient stay (Yes or No) |
| Lactate | Lactate level displayed on blood gas measurement, where available, performed on the day of COVID-19 clinical presentation (mmol/L) |
| Lymphocytes | Lymphocyte count in blood sample obtained by venepuncture, where available, performed on the day of COVID-19 clinical presentation ($10^9/L$) |
| Neutrophils | Neutrophil count in blood sample obtained by venepuncture, where available, performed on the day of COVID-19 clinical presentation ($10^9/L$) |
| Albumin | Minimum albumin across all blood samples obtained by venepuncture, where available, throughout the inpatient stay (g/L) |
| Ferritin | Maximum ferritin across all blood samples obtained by venepuncture, where available, throughout the inpatient stay (ng/L) |
| D-Dimer | Maximum D-Dimer across all blood samples obtained by venepuncture, where available, throughout the inpatient stay (ng/ml) |
| Fibrinogen | Maximum fibrinogen across all blood samples obtained by venepuncture, where available, throughout the inpatient stay (g/L) |
| Troponin | Maximum troponin across all blood samples obtained by venepuncture, where available, throughout the inpatient stay (local units) |
| C-Reactive Protein (CRP) Day 0 | CRP in blood sample obtained by venepuncture, where available, performed on the day of COVID-19 clinical presentation (mg/L) |
| CRP Day 1-2 | Maximum CRP in blood sample obtained by venepuncture performed between days 1 and 2 since the day of COVID-19 clinical presentation, where available (mg/L) |
| CRP Day 3-4 | Maximum CRP in blood sample obtained by venepuncture performed between days 3 and 4 since the day of COVID-19 clinical presentation, where available (mg/L) |
| CRP Day 5-6 | Maximum CRP in blood sample obtained by venepuncture performed between days 5 and 6 since the day of COVID-19 clinical presentation, where available (mg/L) |
| CRP Day 7-8 | Maximum CRP in blood sample obtained by venepuncture performed between days 7 and 8 since the day of COVID-19 clinical presentation, where available (mg/L) |
| CRP Day 9-10 | Maximum CRP in blood sample obtained by venepuncture performed between days 9 and 10 since the day of COVID-19 clinical presentation, where available (mg/L) |
| CRP Day 11-12 | Maximum CRP in blood sample obtained by venepuncture performed between days 11 and 12 since the day of COVID-19 clinical presentation, where available (mg/L) |
| CRP Day 13-14 | Maximum CRP in blood sample obtained by venepuncture performed between days 13 and 14 since the day of COVID-19 clinical presentation, where available (mg/L) |
| CRP Day 15-20 | Maximum CRP in blood sample obtained by venepuncture performed between days 15 and 20 since the day of COVID-19 clinical presentation, where available (mg/L) |

### 2.3 Data Collection

The hospital Picture Archiving and Communication System (PACS) was searched to get the details of the CT scan reports of patients with suspected COVID-19 changes, from 01/03/2020 till 22/04/2020. Reports dictated by a consultant radiologist, and CT scan images where required, were screened for all patients who have changes suggestive of COVID-19 [17]. The radiologically positive cases were included in the study. A record of all the COVID-19 RT-PCR positive swabs was obtained from microbiology department. After removing the duplicates, the CT positive and RT-PCR swab positive cases were populated to a Microsoft Excel spreadsheet. Further patient data from the hospital Electronic Patient Record System (EPR), or eCare, was collected in accordance with data protection and Good Clinical Practise (GCP) guidelines, on a hospital computer, by a team of physicians. Specific instructions were issued to the team of physicians to use during the collection of data to ensure homogenous, standardized interpretation of data from eCare. Healthcare staff who had historically recorded patient information on the EPR during clinical assessment were, of course at the time, blind to the outcomes and hypotheses of this study. All data was checked for systematic error by at least 1 other physician. After data collection, data was fully anonymized.

### 2.4 Independent Predictor Variables

Independent predictor variables were selected for inclusion in this study a priori based on three criteria; (i) having a postulated role for influencing COVID-19 severity based on surrounding literature, (ii) values expected to be available for at least one-third of study participants and (iii) values being collected during the routine care of study participants. 44 independent predictor variables were used for analysis in this study, which are shown below in Table 2.

**Table 2.** The heat-mapped probabilities of inpatient death of patients at different age groups.

| Inpatient Mortality (IPD) | Age Category (Years) |  |  |
| --- | --- | --- | --- |
| | $\leq 40$ | 41 – 69 | $70 \leq$ |
| Yes | 13.97% | 31.50% | 68.30% |
| No | 86.03% | 68.50% | 31.70% |

### 2.5 Outcomes

All patients were either discharged or deceased and thus achieved all four outcomes; (i) inpatient mortality (IPD), (ii) duration of COVID-19 treatment (ADT), and (iii) maximum level of oxygen or ventilatory support during inpatient stay (MOoVS), which was divided into 4 categories: (A) requiring room air or oxygen, (B) requiring high-flow oxygen defined as using a venturi mask, (C) requiring non-invasive ventilation (continuous positive airway pressure (CPAP) or bi-level positive airway pressure (BiPAP)) and (D) requiring intubation. A new confirmed diagnosis of pulmonary embolism during inpatient stay (NCPE) was the fourth outcome. The outcomes were selected for this study a priori based on 3 criteria: (i) All patients will be able to reach one of the pre-defined study outcomes, (ii) The pre-defined study outcomes are representative of COVID-19 severity based on the surrounding literature at the time of study design and (iii) The pre-defined study outcomes will involve data collected in the routine care of study participants. The follow-up period was defined as 2 months to give all patients ample time to achieve a study outcome prior to outcome data collection. During the above-mentioned data collection time-period, if a patient was admitted more than once, and both times were for COVID-19 related reasons within 5 days of each other, this was counted as a failed discharge and thus 1 admission. Days spent in hospital for social reasons or alternative diagnoses prior to developing COVID-19 were subtracted from the duration of inpatient stay to derive the duration of inpatient treatment outcome. The day of COVID-19 clinical presentation was retrospectively determined by physicians during data collection, after careful analysis of the patient notes, in order to determine the first day during the inpatient stay where COVID-19 was diagnosed clinically, based on the full repertoire of available clinical information.

### 2.6 Data Pre-processing

In this study, for a small sample size of n=355 patients, we wish to model the outcomes described in Section 2.5 based on 44 independent variables described in Table 2. However, it was desired to develop a hybrid Bayesian network or other types of probabilistic supervised machine learning methods for modelling continuous and categorical random variables together but learning a suitable model for this messy and small dataset would be almost impractical. As a result, we first convert all the continuous random variables to the categorical variables based on the discretisation method proposed in [18] and taking into account the medical experts’ opinions. In the next stage, we need to deal with the considerable missing values of several variables, which could have a significant impact on the derived results and conclusions made for the health decision makers. The strategies to overcome missing data impact will be discussed in Section 2.7.

#### 2.6.1 Missing data analysis

Observing missing values in a real-world research is quite common to happen due to various reasons [19]. It is of great importance to either appropriately estimate or fill the missing values before a suitable statistical or machine learning method was selected to data, or appropriate adjustment must be made to the selected model to make them robust against the missing values impact [20].

There are various techniques to handle missingness including listwise deletion, pairwise deletion, dummy variable adjustment, marginal mean imputation, regression imputation, maximum likelihood (ML), and multiple imputation (MI) [21,22]. The listwise deletion technique that results in unbiased estimations is recommended by [21] to deal with the missing data mechanism, which depends on the values of the independent variables only. In this study, no outcome (or dependent) variable was missing, but there were several independent variables with high number of missing values. The biochemistry features, including max CRP levels at the different days were among the independent random variables with the highest percentages of missing values ranging from 40% to 70%. However, limited number of missing values could be observed on the rest of independent variables. Due to the high number of missing values in many of the independent random variables, the multiple imputation technique was selected as the most suitable technique and used to estimate the missing values in the dataset.

#### 2.6.2 Balancing Outcomes

As discussed in Section 2.6, we first need to convert all the continuous random variables to the categorical variables based on the discretisation method proposed in [18] and taking into account the medical experts’ opinions. After converting the continuous and multi-scale discrete independent and dependent random variables into the categorical ones, and estimating the missing values using the MI technique (see Section 2.6.1 and [21,22]), it was observed that the resulting outcome variables suffer from a considerable imbalance. For example, only 4.8% of NCPE outcome responses were “Yes” and 95.2% were “NO”. The other outcomes, including IPD, ADT and MOoVS were also severely imbalanced (see Section 3 for the details). The predictions resulted from fitting a suitable ML/Statistical model (e.g., BN) to the imbalanced datasets, whereby one class is dominant, would be inherently biased towards the dominant class, thus decreasing the reliability of the predictions made by the models [23–25]. In supervised machine learning, different methods can be used to address the issues caused by an imbalanced dataset [24,25]. These include: (i) Cost-sensitive learning, which involves manipulating the threshold values, (ii) Imbalanced dataset pre-processing, which can involve oversampling, under-sampling, or Synthetic Minority Oversampling Technique (SMOTE), (iii) Algorithm level approaches, such as active learning and kernel modifications, and (iv) Ensemble learning, such as cost-sensitive boosting [25]. The literature suggests that there is no best strategy to deal with the issues caused by imbalanced datasets [25].

In this study, the authors used the SMOTE to overcome the imbalance in the dataset [26]. The advantage of this technique over other oversampling methods is that it decreases the imbalance in a dataset by synthetically creating new examples of the minority class, and not duplicating them [26,27]. The authors applied the SMOTE on the entire dataset, in concordance with the surrounding literature [28–31]. The details of the balanced outcomes resulted from performing SMOTE are given in Sections 3.1-3.4.

#### 2.6.3 Feature selection

One essential stage in the development of predictive models using supervised Machine Learning (ML) techniques is the selection of relevant variables [23]. This process, which is known as feature selection, includes identifying and choosing the best combination of independent variables in a dataset for efficient and optimum analysis of the problem at hand [23,24]. The rationale behind it is that in a dataset, some variables can be redundant, for example due to multicollinearity, or not relevant to the response. The presence of redundant and irrelevant features in a dataset can seriously hamper the accuracy of the predictions. By performing robust feature selection, predictive models that perform optimally on both seen and unseen data could be developed effectively. Hence, feature selection attempts to discard irrelevant and redundant independent variables that do not contribute to the development of a reliable predictive model [25]. It is noteworthy that in the process of selecting variables, only the training data are considered to avoid inaccurate estimates of the test errors [25,26].

There are three methods for selecting a subset of features [27,28]. Filter methods use statistical properties of the features, such as correlation coefficients, F-test, T-test, or information-theory based measures, such as mutual information and interaction information, in order to rank features based on their relevance to the response and other variables [23,27,29,30]. These methods can be grouped into univariate filter methods and multivariate filter methods. Univariate filter methods rank features only based on their relevance to the response, whereas multivariate filter methods consider the interaction between features as well [27].

Wrappers are the second method for feature selection [23]. In this group of techniques, a machine learning model is used to score subsets of features based on the predictive power of the method. The process of feature selection can be categorized into forward-selection, backward-elimination, and mixed selection [26]. Forward-selection methods start modelling with zero predictors (a base model), choose features step-by-step, and evaluate the performance. Whereas, backward feature elimination methods start with the complete set of independent variables and look for an optimum subset of variables with the best performance through stepwise elimination of non-informative features [26]. Mixed methods combine forward-selection and backward elimination techniques. Wrappers use cross-validation to optimise the performance of the learning method in order to select the optimum subset of variables [27].

A third approach that is sometimes grouped with wrappers [23] is the embedded or intrinsic method [27,31]. Similar to wrappers that select a subset of variables based on a learning model, embedded methods embed this process into its predictive model development [27]. For instance, if the variable importance measure of the random forest method is used to improve the performance of a random forest model, then it is an embedded method. Whereas, if this capability is used to select features and develop predictive models with methods other than the random forest, then it is a wrapper method.

For its feature selection, this study adopts the recursive feature elimination (RFE) method, which is a backward variable selection wrapper technique [31]. For this purpose, the authors computed the RFE method in R (version 4.0.2) using the random forest (RF) function embedded in the Caret package [32,33]. As the first stage, the dataset is randomly split into 70% training and 30% testing using the validation set approach [26]. Subsequently, using the training data, a predictive model containing all features is developed based on the random forest method. This model then ranks the features based on a measure of importance. Consequently, the RFE method eliminates the least important feature, develops a new model based on a smaller number of independent variables, and re-ranks the remaining predictors [31]. RFE identifies two parameters: the number of subsets to evaluate and the number of predictors in each of the subsets. For each subset, the process of eliminating the least-important features continues until it reaches a determined subset size. Eventually, RFE compares the predictive performance of all subsets and determines the best subset size with the best accuracy [31]. In this study, the performance of the wrappers is assessed using *k*-fold Cross-Validation (k=10), which repeats five times.

Since the RFE uses a supervised Machine Learning method to perform feature selection, it is essential to evaluate the performance of the resulting RF model. In supervised ML, the validation set approach, *k*-fold Cross-Validation (kfCV), and Leave-One-Out Cross-Validation (LOOCV) are the three main methods to perform this task [26]. The kfCV method randomly divides the dataset into *k* groups of observations (folds) with roughly equal sizes. The kfCV method uses the first fold for testing the model and the remaining *k*-1 folds for training it. This process repeats *k* times until all folds are once used for testing the performance of the model developed with the remaining *k*-1 folds. These *k* performances are averaged to calculate the mean performance of the predictive model [26]. While *k* is an arbitrary number and can take any number less than the number of observations in a dataset, empirically, values of 5 or 10 show resistance against high bias or variance [26]. In this paper, *k*=10 guarantees a higher number of training observations in each fold to improve the predictive performance of the model [26].

The kfCV is advantageous over the validation set approach and LOOCV for the following reasons. In the case of the former, since the dataset is randomly divided into a training and a testing set, if the process repeats, it could result in different predictions. And in the case of the latter, the LOOCV tends to result in higher variance on the unseen observations [26].

The result of the feature selection using the RFE method for each of these responses is shown in Table 3. Moreover, Figure 1 shows the performance of the RFE method based on the ranks of the variables. In this figure, the red circle shows the maximum achievable performance based on the best combination of variables. For instance, in panel (a) (NCPE outcome); the accuracy of the model using only ‘UoB’ is approximately 81%. By adding variables based on their rank (Table 3), the accuracy of the model improves. In the case of NCPE (panel (a)), after adding “Age”, the accuracy increases to 84%, and so on (Figure 1).

**Table 3:** Feature selection results for four different outcomes, including IPD, ADT, NCPE and MOoVS.

| Predictor | RFE<br>(Response<br>= NCPE) | RFE<br>(Response<br>= MOoVS) | RFE<br>(Response<br>= ADT) | RFE<br>(Response<br>= IPD) |
| --- | --- | --- | --- | --- |
| Age | 2 | 1 | 7 | 1 |
| Gender (Ge) | 7 | 17 | 17 | 33 |
| Ethnicity | 17 | 5 | 24 | 4 |
| Oxygen Saturations (OS) | 16 | 2 | 9 | 2 |
| Respiratory Rate (BPM) | 19 | 12 | 26 | 9 |
| Temperature | 6 | 10 | 19 | 35 |
| Obesity | 11 | 8 | 11 | 28 |
| Previous Venous Thromboembolism (PVTE) | Rejected | 33 | 33 | 36 |
| Chronic Obstructive Pulmonary Disease (COPD) | Rejected | 37 | 37 | 37 |
| Bronchiectasis | Rejected |  | 41 | 31 |
| Asthma | 21 | 27 | 34 | 10 |
| Interstitial Lung Disease (ILD) | Rejected | 21 | 40 | 38 |
| Lung Cancer (LC) | Rejected | 41 | 38 | 39 |
| Diabetes Mellitus (DM) | 29 | 16 | 27 | 21 |
| Hypertension (HTN) | 14 | 26 | 8 | 25 |
| Ischaemic Heart Disease (IHD) | 31 | 28 | 28 | 16 |
| Chronic Kidney Disease (CKD) | 32 | 31 | 31 | 8 |
| Non-steroidal anti-inflammatory drugs (ANNC) | 33 | 38 | 25 | 15 |
| Anticoagulant | 23 | 35 | 29 | 17 |
| Long-Term Antibiotic (LTA) | Rejected | 34 | 36 | 30 |

| **Predictor** | **RFE**<br>**(Response**<br>**= NCPE)** | **RFE**<br>**(Response**<br>**= MOoVS)** | **RFE**<br>**(Response**<br>**= ADT)** | **RFE**<br>**(Response**<br>**= IPD)** |
| --- | --- | --- | --- | --- |
| Long Term Oral Steroid (LTO) | Rejected | 39 | 42 | 42 |
| Immunosuppressants (ISES) | Rejected | 32 | 39 | 32 |
| Oral NSAIDs (ONS) | Rejected | 40 | 32 | 41 |
| Angiotensin Converting Enzyme Inhibitors (ACEI) | 28 | 36 | 30 | 40 |
| Angiotensin Receptor Blockers (ARB) | 27 | 29 | 35 | 27 |
| CT imaging severity of COVID-19 related changes (UoB) | 1 | 4 | 2 | 23 |
| COVID-19 related Chest X-Ray changes (CCX) | 30 | 7 | 10 | 11 |
| Lactate (LDP) | 12 | 25 | 21 | 20 |
| Lymphocytes (LyDP) | 4 | 23 | 16 | 18 |
| Neutrophils (NDP) | 5 | 18 | 15 | 26 |
| Albumin (MADA) | 3 | 6 | 1 | 6 |
| Ferritin | 24 | 20 | 23 | 24 |
| D-Dimer (MDD) | 8 | 11 | 6 | 7 |
| C-Reactive Protein (CRP) Day 0 | 18 | 13 | 3 | 19 |
| CRP Day 1-2 (MCRP1) | 13 | 22 | 4 | 12 |
| CRP Day 3-4 (MCRP3) | 20 | 19 | 14 | 22 |
| CRP Day 5-6 (MCRP5) | 10 | 15 | 12 | 5 |
| CRP Day 7-8 (MCRP7) | 9 | 3 | 5 | 3 |
| CRP Day 9-10 (MCRP9) | 22 | 14 | 13 | 14 |
| CRP Day 11-12 (MCRP11) | 26 | 9 | 18 | 29 |
| CRP Day 13-14 (MCRP13) | 15 | 24 | 20 | 34 |
| CRP Day 15-20 (MCRP15) | 25 | 30 | 22 | 13 |

**Table 3.**
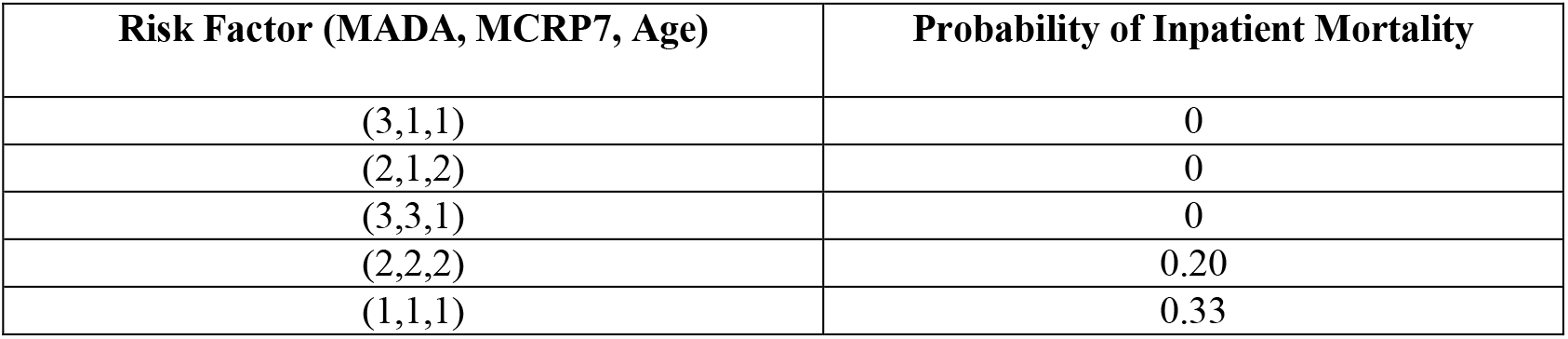
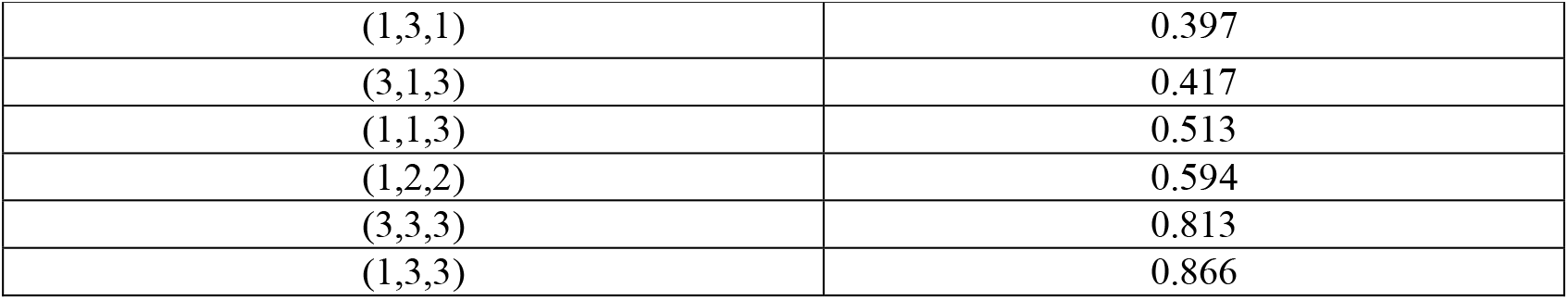
The conditional probability of IPD given different configurations of the parent nodes. MADA (1= ‘≤30’, 2= ‘30-35’, 3= ‘35≤‘), MCRP7 (1= ‘≤50’, 2= ‘51-100’, 3= ‘100≤‘), and Age in years (1= ‘≤40’, 2= ‘40-70’, 3= ‘70<‘).

**Figure 1.**
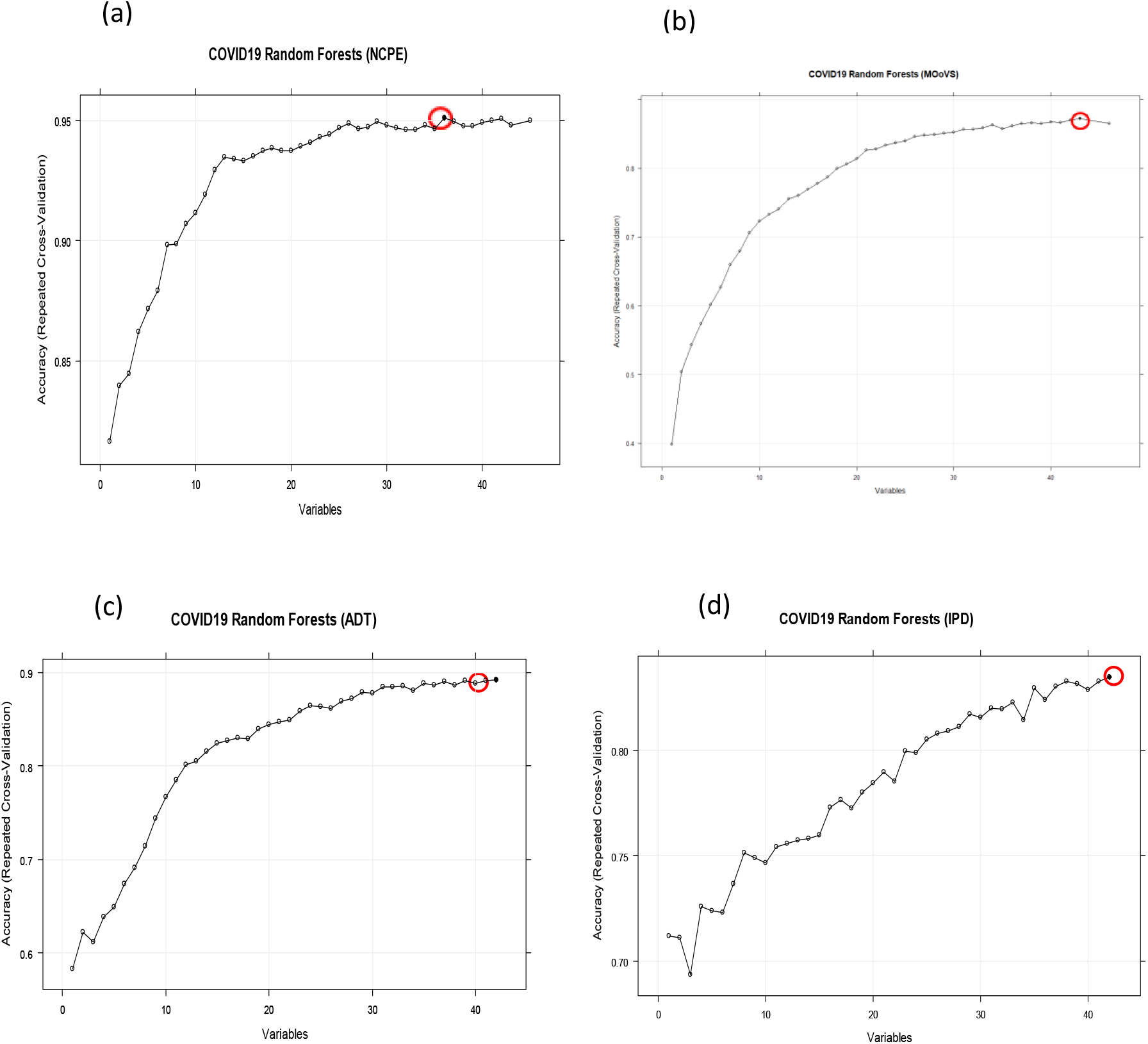
a-d: Performance of the RFE based on the ranks of the features a) NCPE, b) MOoVS, c) ADT and d) IPD. The red circle shows the maximum achieved accuracy.

Table 3 and Figure 1 (a)-(d) show that for each outcome, there is a specific combination of independent variables that produce the highest predictive performance among all other possible combination of variables for the selected outcome. For instance, UoB, Age, MADA, LyDP, NDP, temperature, gender, D-Dimer (MDD), MCRP7, MCRP5, obesity and LDP, covering 93% accuracy, would be sufficient to model the NCPE outcome. Therefore, these variables will be used for developing the probabilistic predictive model. The numbers against each variable for the corresponding response indicates the predictive importance of that factor. The details of the most relevant variables affecting each response variable will be provided in Section 3.

### 2.7 Bayesian Networks

A Bayesian network (BN) is a probabilistic graphical model that is used to represent knowledge about an uncertain domain [34]. Each random variable is represented by a node in the BN. A conditional probability table (CPT) is attached to each node. A link, or ‘edge’, between two nodes represents a probabilistic dependency between the linked nodes. The ‘directed’ links are shown with an arrow pointing from the causal node to the effect node. There must not be any directed cycles: one cannot return to a node simply by following a series of directed links. This means that BNs are Directed Acyclic Graphs (DAGs). Nodes without a child node are called leaf nodes, nodes without a parent node are called root nodes, and nodes with parent and child nodes are called intermediate nodes. A BN represents dependence and conditional independence relationships among the nodes using joint probability distributions, with an ability to incorporate human oriented qualitative inputs. The method is well established for representing cause-effect relationships.

Applications of BN methods are found in a growing number of disciplines and policies [35,55]. BNs are particularly useful for evaluation because of their capability of classification based on observations. Moreover, a BN can do unsupervised learning from a dataset and allow the learning algorithm to find both structure and probabilities. This means the policy-, or decision-maker does not need to know how to learn the BN, although it is possible to aid the learning algorithm with a priori knowledge about relations and probabilities. Dealing with uncertainty when evaluating policy is a challenge that can be addressed using BNs because some uncertain probabilities of variables may be safely ignored to get to the desired probabilistic quantity of a random variable. Furthermore, BNs engage directly with subjective data in a transparent way. Hence, the method could be considered as a tool to explore beliefs, evidence, and their logical implications, as opposed to a means of proving concepts in some absolute sense.

BN learning consists of two general steps:

- Finding DAG (or the network structure) which illustrates the inter-dependency between the variables/nodes, and denoted by *G*:
- Finding CPT (or conditional probability density) for each node given the values of its parents on the learned network structure *G*.

Finding the best DAG is the crucial step in BN design. Construction of a graph to describe a BN is commonly achieved based on probabilistic methods, which utilize databases of records [29,36,46, 54], such as the search and score approach. In this approach, a search through the space of possible DAGs is performed to find the best DAG. The number of DAGs, *f*(*p*), as a function of the number of nodes, *p*, grows exponentially with *p* [37].

## 3. Results

### 3.1 Modelling inpatient mortality (IPD) using BNs

In this section, we provide the details of modelling inpatient mortality (IPD) using BN as described in the methods. Modelling the IPD based on the massively incomplete and imbalanced data using BN models is very challenging. Prior to learning the BN structure and corresponding CPTs, the original categorised data was completed by estimating the missing values of the independent variables using the multiple imputation technique (see also Section 2.6.1). We then used the ML-based technique known as “SMOTE” which is briefly explained in Subsection 2.6.2, to balance data with respect to “IPD” response variable. It should be noted that 30% of IPD outcome responses were “Yes” and 70% were “NO”. We then perform the feature selection approaches as described in Section 2.6.3, to identify the most important risk factors affecting the inpatient mortality (IPD). The results of the feature selection are reported in Table 3, Figure 1, and Figure 14 in appendix A.

The BN structure learned from the data only for IPD based on the most important factors affecting IPD (as illustrated in Table 3) is shown in Figure 2. This network structure was learned from the completed data by evaluating the best model out of various score-based or constraints-based methods [38]. From this model, it is evident that the way that several independent variables affecting IPD is not correct, and it should be in reverse. For example, Age, MADA and OS are among the most important risk factors, which affect IPD rather than be affected by IPD. Therefore, it was important to discuss the resulting BN model, illustrated in Figure 2 with the domain medical experts, and consequently revise this BN by considering experts’ opinions. The revised BN model learned based on the combination of data and expert opinions, whilst also validated using several model diagnostic algorithms, such as *k*-fold cross validation, is illustrated in Figure 3.

**Figure 2.**
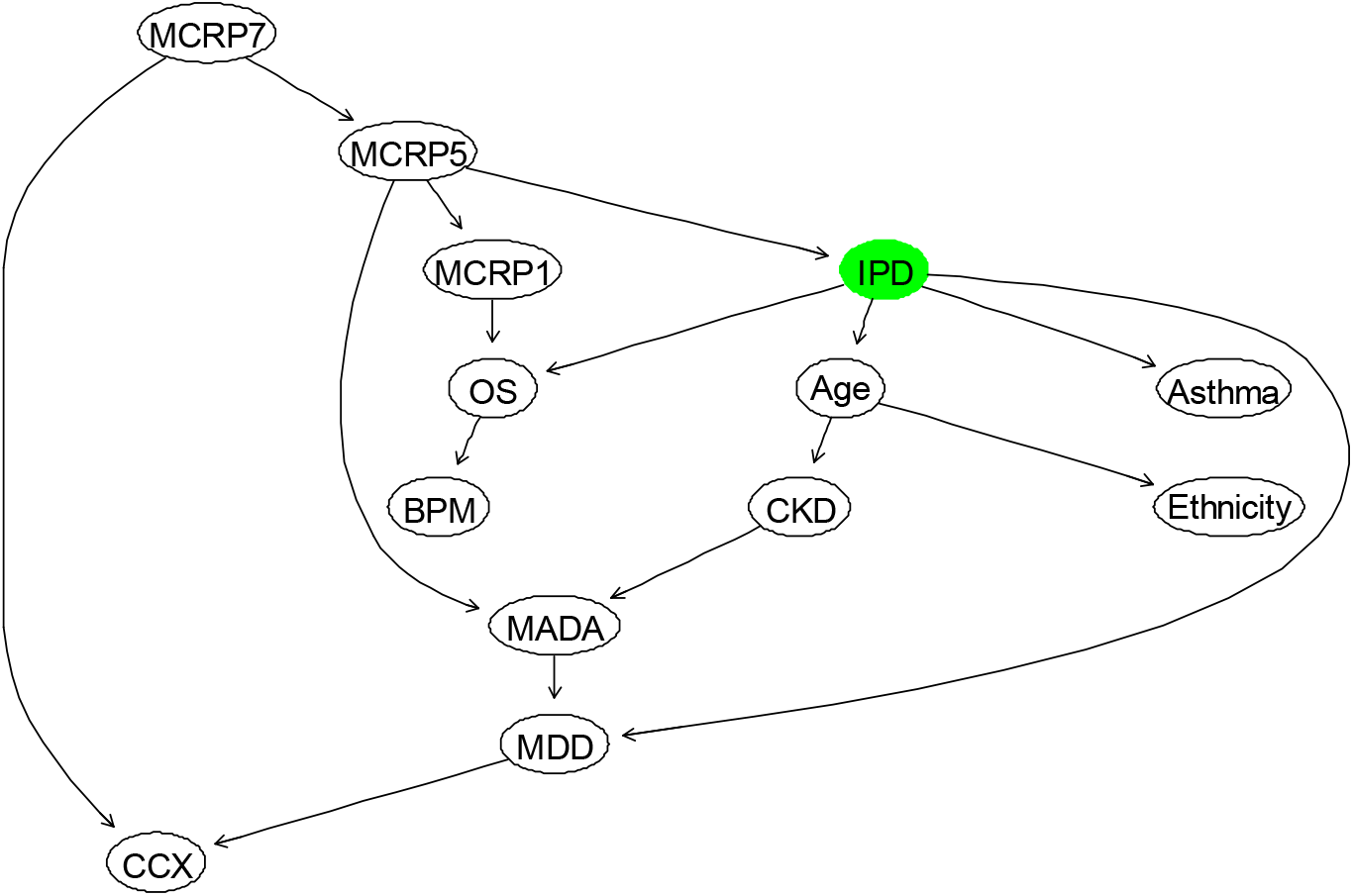
The BN that is fully learned from data to model “IPD” in terms of other relevant factors.

**Figure 3.**
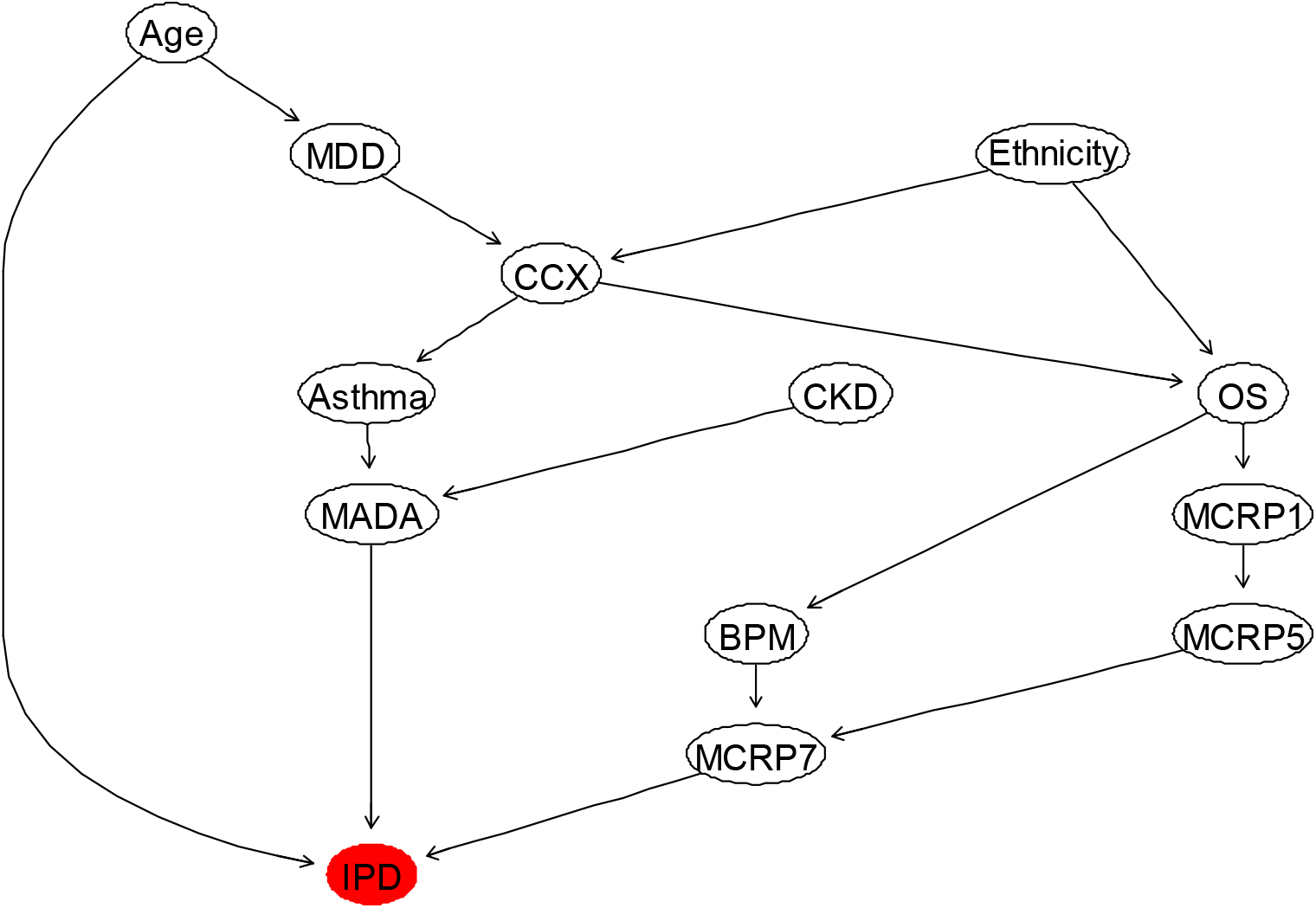
The BN learned by eliciting the domain expert combined with the (balanced and completed) data.

**Figure 3.**
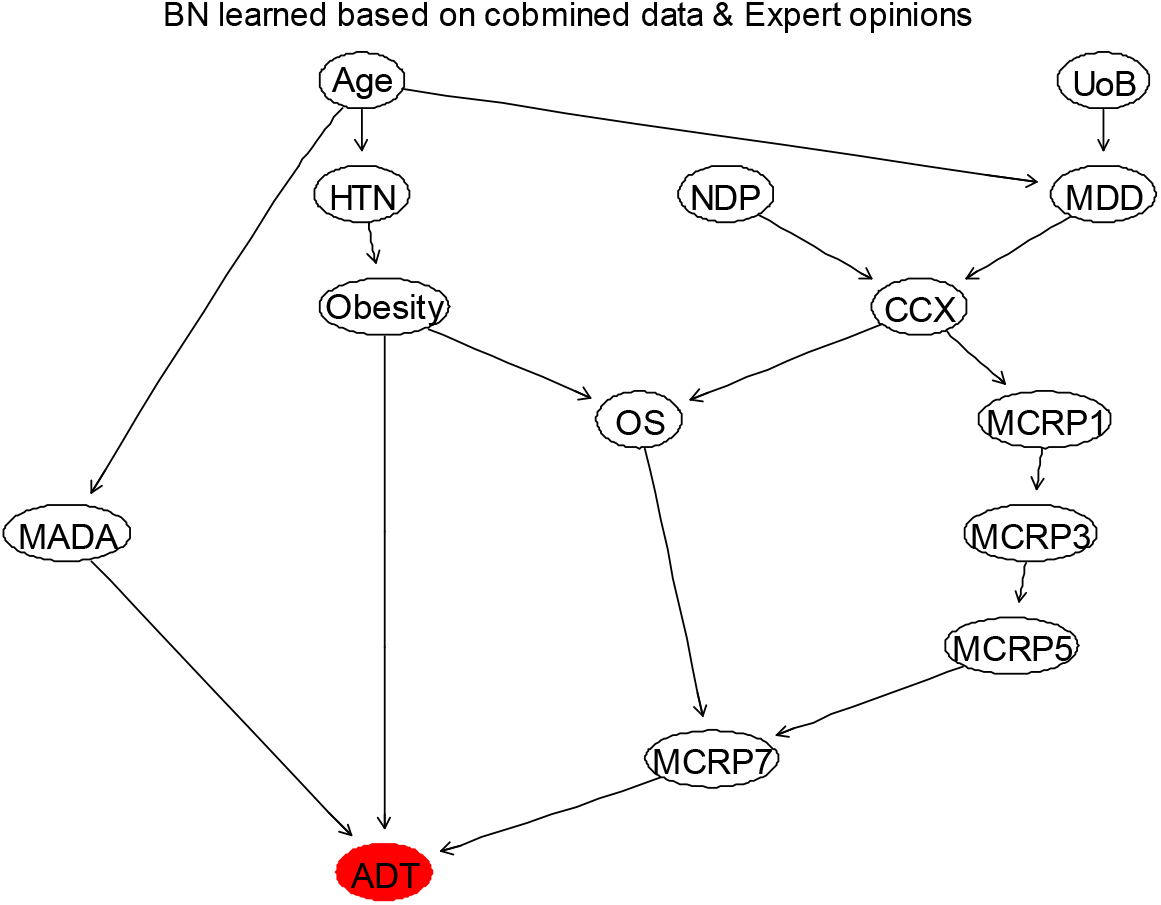
The BN that is learned from the combination of data and expert opinions, to model “ADT” in terms of the first 11 most relevant factors.

In the BN model proposed for modelling IPD, the strength of the link, as well as the associated uncertainty, is captured using probabilities and statistical distributions, which are estimated or derived based on the observed data. Figure 4 shows the learned BN with the estimated marginal probabilities shown on each node. In this BN, three nodes (Age, CKD, and Ethnicity) are considered as root nodes, and their parameters are learned by estimating these probabilities using the maximum likelihood method (MLE) or Bayes estimate. The estimated marginal and conditional probabilities for the variables can be updated in the light of new evidence or data using a statistical algorithm known as the Bayes rule [36]. Hence, the BN can compute the probability of surviving or dying due to COVID-19, based on the different combination of the parent nodes, including Age, the minimum Albumin level during admission (MADA), and the mean CRP level during days 7-8 since clinical presentation of COVID-19 (MCRP7). For example, Table 4 shows the probabilities of death and survival of patients at different age groups. It confirms the reported findings that as the age of a patient increases; the risk of mortality also increases. To be precise, the death rate of Covid-19 patients’ ≥70 years is 5 times larger than patients’ ≤40 years. These probabilities can be updated by observing more evidence about the states of other influencing variables.

**Figure 4.**
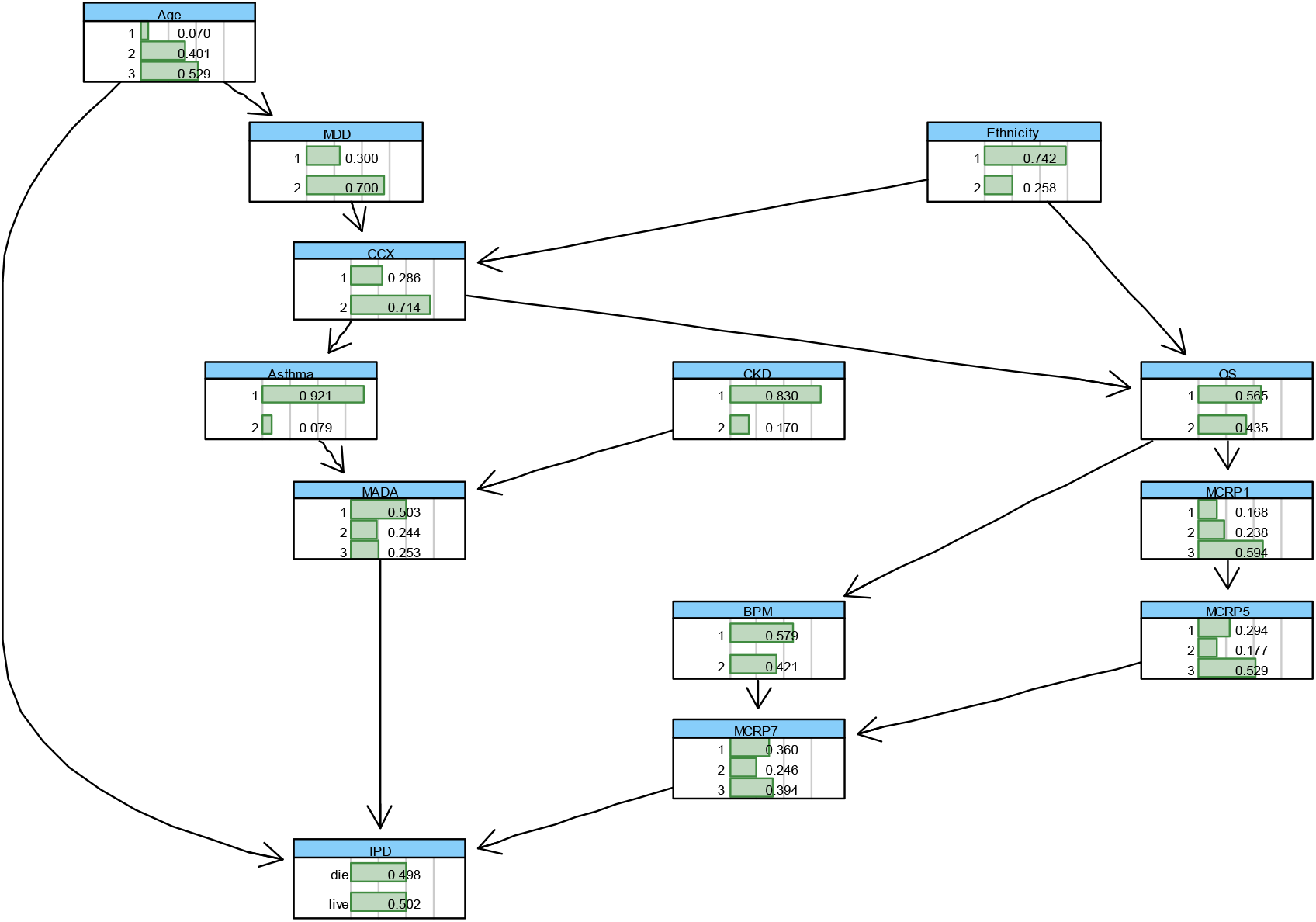
The BN with conditional probability tables learned for “IPD” outcome based on the combined elicited domain expert opinions with the (balanced) data.

**Figure 4.**
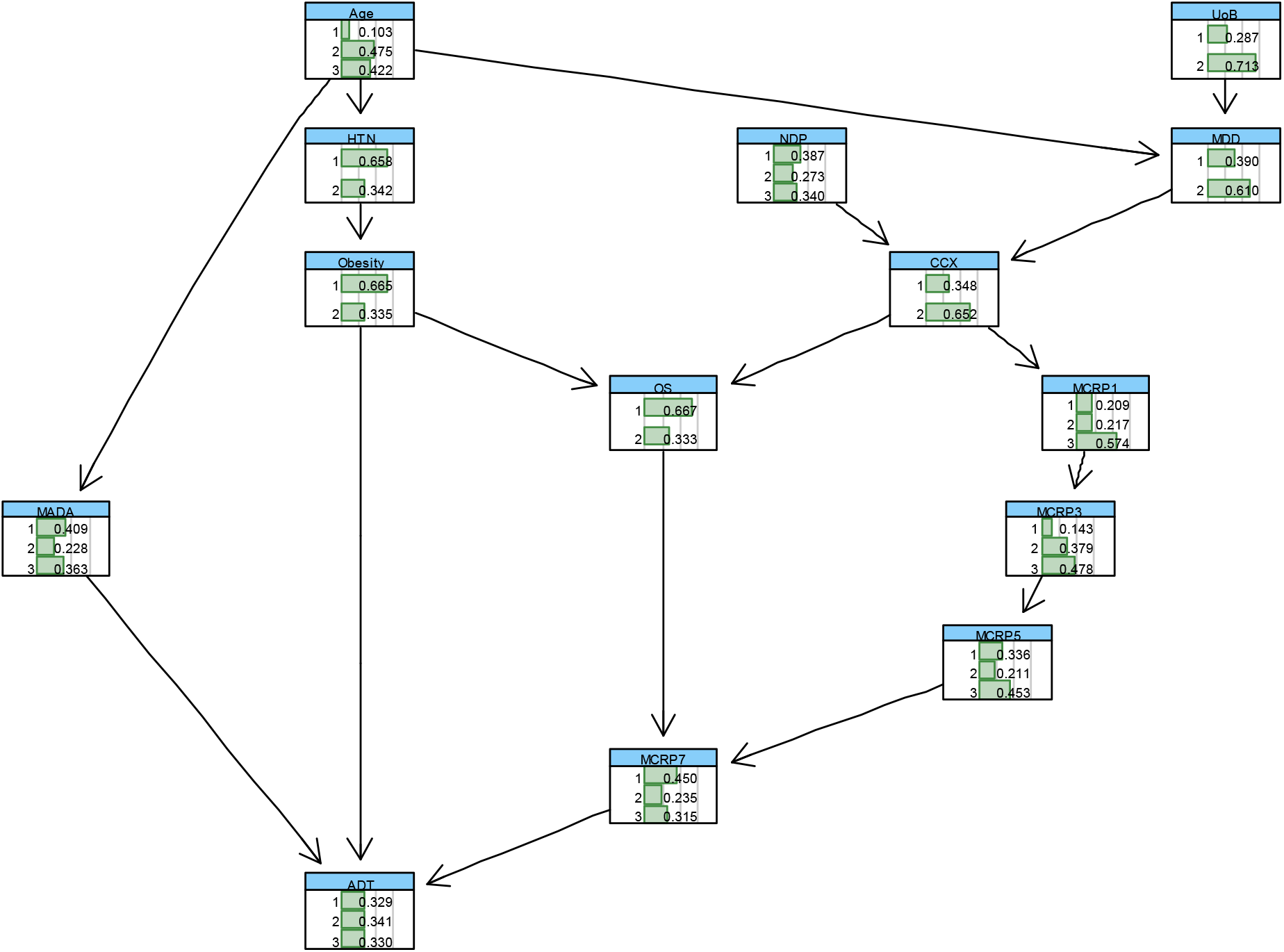
The BN with conditional probability tables learned for “ADT” outcome based on the combined elicited domain expert opinions with the (balanced) data.

**Table 4.**
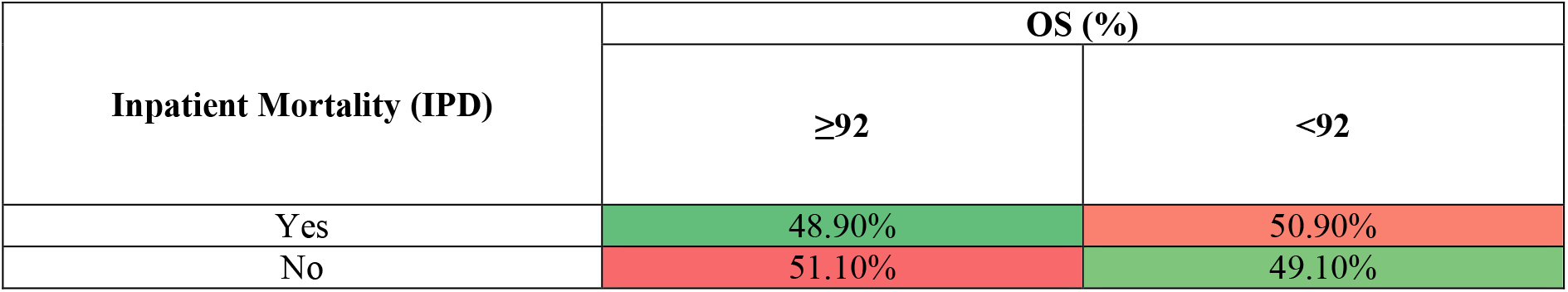
The heat-mapped probabilities of death and survival of patients based on OS.

Table 5 shows the conditional probabilities of IPD given the selected configurations of its parent nodes, including Age, MADA and MCRP7. It can be concluded that for the patients in the first age group (≤40 years) if the MADA is above 35 g/L, they will survive the Covid-19 regardless of MCRP7. For a patient in this age group and with the MADA level less than 30 g/L, they would more likely survive if their MCRP7 were less than 50 mg/L. This means that when the MADA level is less than 30 g/L, reflective of malnourishment and frailty, patients are at a particularly high risk especially if their MCRP7 level is above 50 mg/L. Interestingly, similar patterns can be found for the patients aged ≥70 years old, but the corresponding survival probabilities are considerably lower. Overall, these results indicate how albumin negatively, whilst CRP and age positively, correlate with inpatient mortality in an additive manner.

**Table 5.**
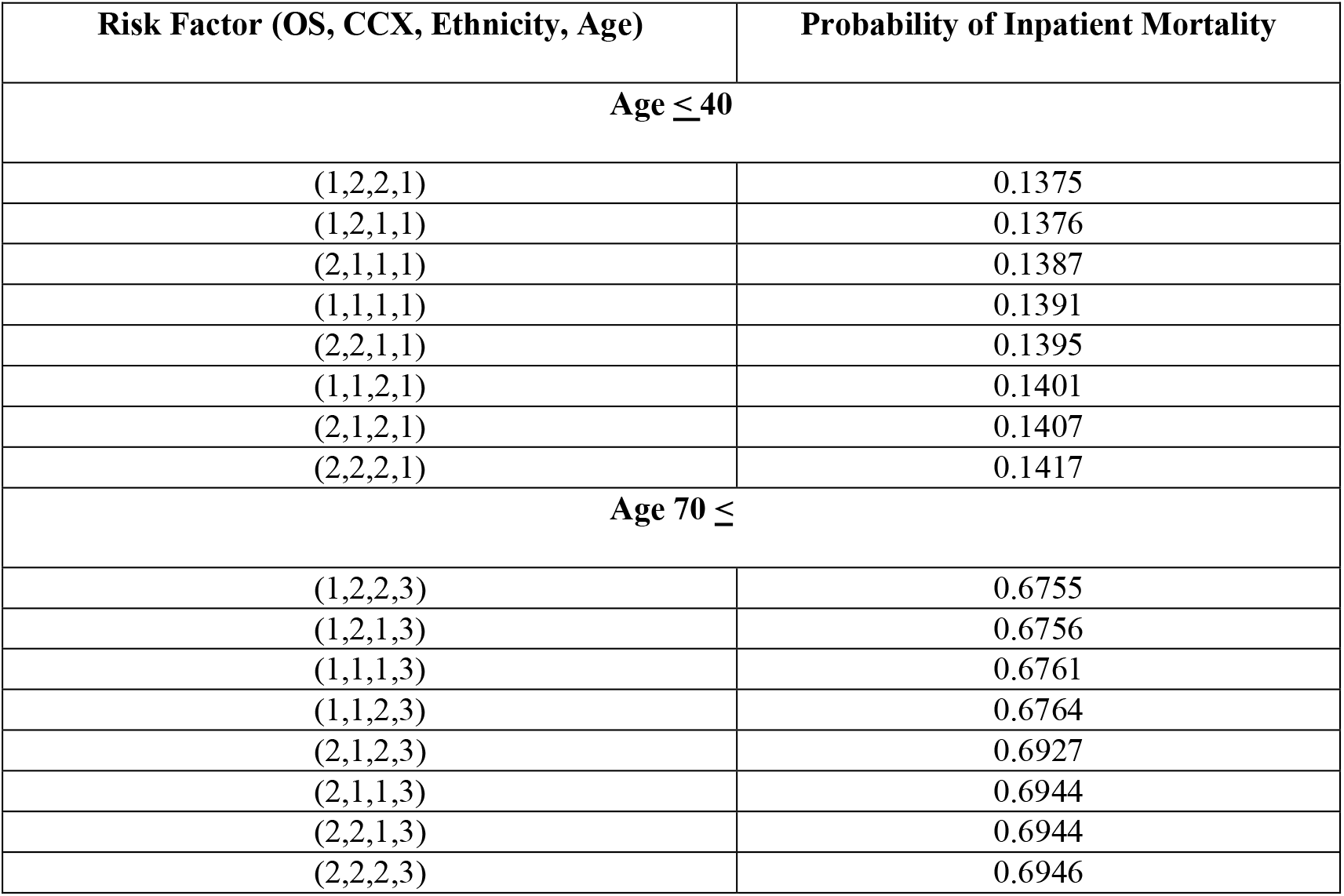
The conditional probability of IPD given different configurations of Oxygen Saturations (OS), Ethnicity, Changes on Chest X-ray (CCX) and Age. OS in % (2= ‘<91’, 1= ‘92≤‘), CCX (1= ‘No’, 2= ‘Yes’), Ethnicity (1= ‘Caucasian’, 2= ‘Non-Caucasian’), and Age in years (1= ‘≤40’, 3= ‘70≤‘)

| Risk Factor (OS, CCX, Ethnicity, Age) | Probability of Inpatient Mortality |
| --- | --- |
| **Age $\leq 40$** |  |
| (1,2,2,1) | 0.1375 |
| (1,2,1,1) | 0.1376 |
| (2,1,1,1) | 0.1387 |
| (1,1,1,1) | 0.1391 |
| (2,2,1,1) | 0.1395 |
| (1,1,2,1) | 0.1401 |
| (2,1,2,1) | 0.1407 |
| (2,2,2,1) | 0.1417 |
| **Age $70 \leq$** |  |
| (1,2,2,3) | 0.6755 |
| (1,2,1,3) | 0.6756 |
| (1,1,1,3) | 0.6761 |
| (1,1,2,3) | 0.6764 |
| (2,1,2,3) | 0.6927 |
| (2,1,1,3) | 0.6944 |
| (2,2,1,3) | 0.6944 |
| (2,2,2,3) | 0.6946 |

Table 6 shows the probabilities of IPD given the lowest recorded oxygen saturation (OS) on the day of clinical presentation with COVID-19, computed from the learned BN model illustrated in Figure 4. As OS decreases, the risk of mortality increases, suggesting that there is a negative association between these two variables.

**Table 6.** The conditional probability of IPD given different configurations of MCRP1 (1= ‘≤30’, 2= ‘31-100’, 3= ‘100≤‘) and MCRP7 (1= ‘≤50’, 2= ‘50-100’, 3= ‘100≤‘), Age in years (1= ‘≤40’, 3= ‘70≤‘) and MADA (1= ‘≤30’, 2= ‘30-35’, 3= ‘35≤‘). *denotes a combinations of risk factors whereby CRP levels are increasing, between days 1-2 and days 7-8, since clinical COVID-19 presentation.

| Risk factor (MADA, Age, MCRP1, MCRP7) | Probability of Inpatient Mortality |
| --- | --- |
| **MADA $\geq 35$ & Age $\leq 40$ years** |  |
| (3,1,2,1) | 0 |
| (3,1,3,1) | 0 |
| (3,1,1,2) * | 0 |
| (3,1,1,3) * | 0 |
| **MADA $\geq 35$ & Age $70 \leq$ years** |  |
| (3,3,2,1) | 0.418 |
| (3,3,3,1) | 0.416 |
| (3,3,1,2) * | 0.496 |
| (3,3,1,3) * | 0.812 |
| **MADA $\leq 30$ & Age $70 \leq$ years** |  |
| (1,3,2,1) | 0.515 |
| (1,3,3,1) | 0.513 |
| (1,3,1,2) * | 0.734 |
| (1,3,1,3) * | 0.865 |

Although the above findings are useful, it is also fascinating to observe how OS could jointly, with other influencing independent variables, affect the risk of inpatient mortality, as shown below in Table 7.

**Table 7.** Summary of the predictive performance results of the BN model developed to model IPD as Illustrated in Figure 4.

| Predictive performance metric | PPV | NPV | Specificity | Sensitivity | Overall accuracy | F1-Score |
| --- | --- | --- | --- | --- | --- | --- |
| BN for IPD | 82% | 67.86% | 82.6% | 85.7% | 84.1% | 83.7% |

Table 7 illustrates the conditional probability of IPD given different configurations of OS, Ethnicity, changes on chest X-ray (CCX), and age of patients. The illustrated results in this table suggest that the risk of inpatient mortality is elevated for patients with reduced oxygen saturations and older patients. Ethnicity seems to increase the risk of death in patients 70≤ years, which is concordant with surrounding literature [4], however the results were not true for younger patients. This may have been due to a skewed population demographic whereby older patients tended to be Caucasian and younger patients reflected a more multicultural demographic. CCX did not seem to significantly affect the risk of IPD, perhaps because the presence of changes is more likely an indicator of the time-point that an individual is along their COVID-19 infection rather than an indicator of severity.

The next important research question is that how the trend in CRP levels at the different days, in addition to the individual CRP levels, can be incorporated and evaluated using an appropriate model. This is because CRP levels can often correlate with infection severity, with a small associated lag time, and therefore the trend in CRP is useful clinically for predicting what will happen to the patient. For example, if the gradient between the latest two CRP variables were positive, it would indicate that the infection is getting worse, whereas if the gradient were negative, it would indicate infection resolution. To account for the gradient between the CRPs, a dynamic version of BN needs to be developed, which is not possible due to the lack of training data. However, the BN model illustrated in Figure 4 can be used to compute the risk of inpatient mortality for different levels of CRP at the different days during the clinical course of COVID-19 infection, as shown below in Table 8.

**Table 8.**
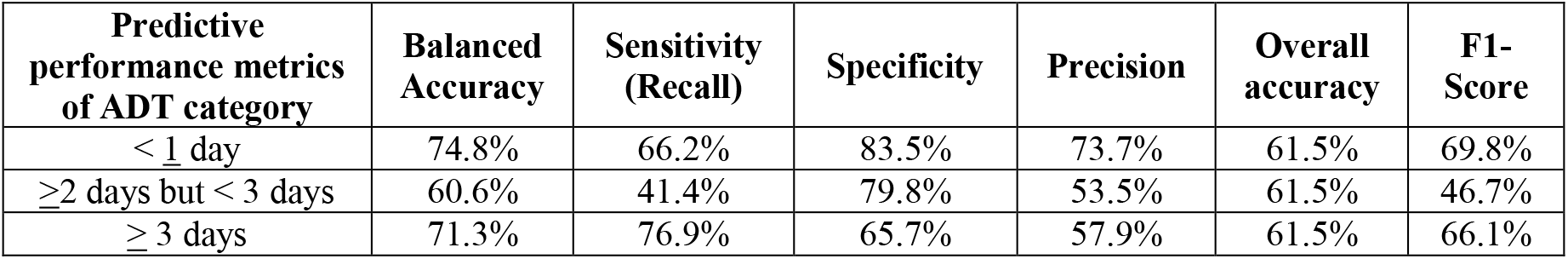
Summary of the predictive performance results of BN model developed to model ADT as Illustrated in Figure 8. The ADT categories are ‘1’ (< <u>1</u> day), ‘2’ (≥2 days but < 3 days) and ‘3’ (≥ 3 days)

Table 8 provides the probabilities of inpatient mortality for the selected configurations of the mean CRP on days 1-2 (MCRP1) and 7-8 (MCRP7) since clinical presentation with COVID-19, as well as Age and MADA. As shown, if the level of MADA is 35≤, increases or decreases in CRP levels during the clinical course of COVID-19 infection will not impose a mortality risk in patients ≤40 years. However, in patients aged 70≤ years, any increase in CRP levels (mg/L) between days 1-2 and days 7-8, would significantly increase mortality risk. In this age group, if MADA levels are also below 30 g/L, and their MCRP increases from ≤30 mg/L, during days 1-2 since clinical COVID-19 presentation, to 100≤ mg/L during days 7-8 since clinical COVID-19 presentation, our data in Table 8 indicates that the risk of inpatient mortality can increase as high as 86.5%.

Table 9 below indicates the Positive predictive value (PPV), Negative predictive value (NPV), Sensitivity, Specificity, Overall accuracy and F-Score, which are used to evaluate the predictive performance of the BN suggested to model IPD in Figure 4, in terms of the feature selected risk factors. James et al., describe the definitions and details of how these metrics can be computed [26].

**Table 9.** The heat-mapped, conditional probabilities of Duration of Treatment for COVID-19 (ADT). The ADT categories are ‘1’ (< <u>1</u> day), ‘2’ (≥2 days but < 3 days) and ‘3’ (≥ 3 days). MCRP1 has been divided into 3 categories: ‘1’ (≤ 50), ‘2’ (51-100) and ‘3’ (≥100). MADA has been divided into 3 categories: ‘1’ (≤ 30), ‘2’ (30-35) and ‘3’ (≥ 35).

| Probability of ADT category given category of Obesity, MADA & MCRP7 | MADA (3) & MCRP1 (1) | MADA (3) & MCRP1 (3) | MADA (1) & MCRP1 (1) | MADA (1) & MCRP1 (3) |
| --- | --- | --- | --- | --- |
| **BMI $< 30$ (Non-Obese patients)** |  |  |  |  |
| $< 1$ day | 71.2% | 68.7% | 10.5% | 10.5% |
| $\geq 2$ days but $< 3$ days | 23.7% | 25% | 32.7% | 30.3% |
| $\geq 3$ days | 5.1% | 6.3% | 56.8% | 59.2% |
| **BMI $\geq 30$ (Obese patients)** |  |  |  |  |
| $< 1$ day | 54.4% | 49.4% | 13.2% | 11% |
| $\geq 2$ days but $< 3$ days | 40.7% | 45.2% | 34.6% | 32.5% |
| $\geq 3$ days | 4.8% | 5.4% | 52.2% | 56.5% |

F1 Score is the Harmonic Mean between precision and recall and Table 9 indicates the high F1 score (83.7%) and accuracy (84.1) of our BN model developed for IPD, despite the small dataset used to train and test our BN model. 82% (PPV) of adult patients predicted to die as inpatients during clinical COVID-19 infection, by our model, will die. However, only 67.86% (NPV) of adult COVID-19 patients predicted to survive the inpatient admission will indeed survive. This indicates that our model may fail to predict inpatient death a sub-set of adult COVID-19 patients, but we expect this to improve with a larger dataset, which also incorporates more variables such as socioeconomic factors.

### 3.2 Modelling Duration of Treatment for COVID-19 using BNs

The next outcome of interest is the duration of treatment for Covid-19 in days (ADT). Like the IPD variable, ADT outcome data was imbalanced. Out of the 355 patients in this study, 100, 88, and 167 patients were admitted to hospital for up to one day (‘1’), 2 to 3 days (‘2’) and 4 days or more (‘3’), respectively. As a result, the dataset was first balanced in terms of the ADT variable, using the methods described in Section 2.61. We then conducted the feature selection, as per methods described in Section 2.62, to identify the most important risk factors affecting ADT. The results of the feature selection are reported in Table 3, Figure 1, and Figure 15 in appendix A. The first 11 most relevant independent variables influencing ADT were then selected, based on the feature selection results to learn a BN for ADT based on the observed processed data. This was then combined with the elicited expert opinions about the network structure. The BN model learned based on the combination of data and expert opinions, and also critically validated against other suitable BNs learned based on the observed data only, using several model diagnostic algorithms, including *k*-fold cross validation, is shown in Figure 8. This illustrates interdependencies between ADT and the most relevant independent variables.

**Figure 5.**
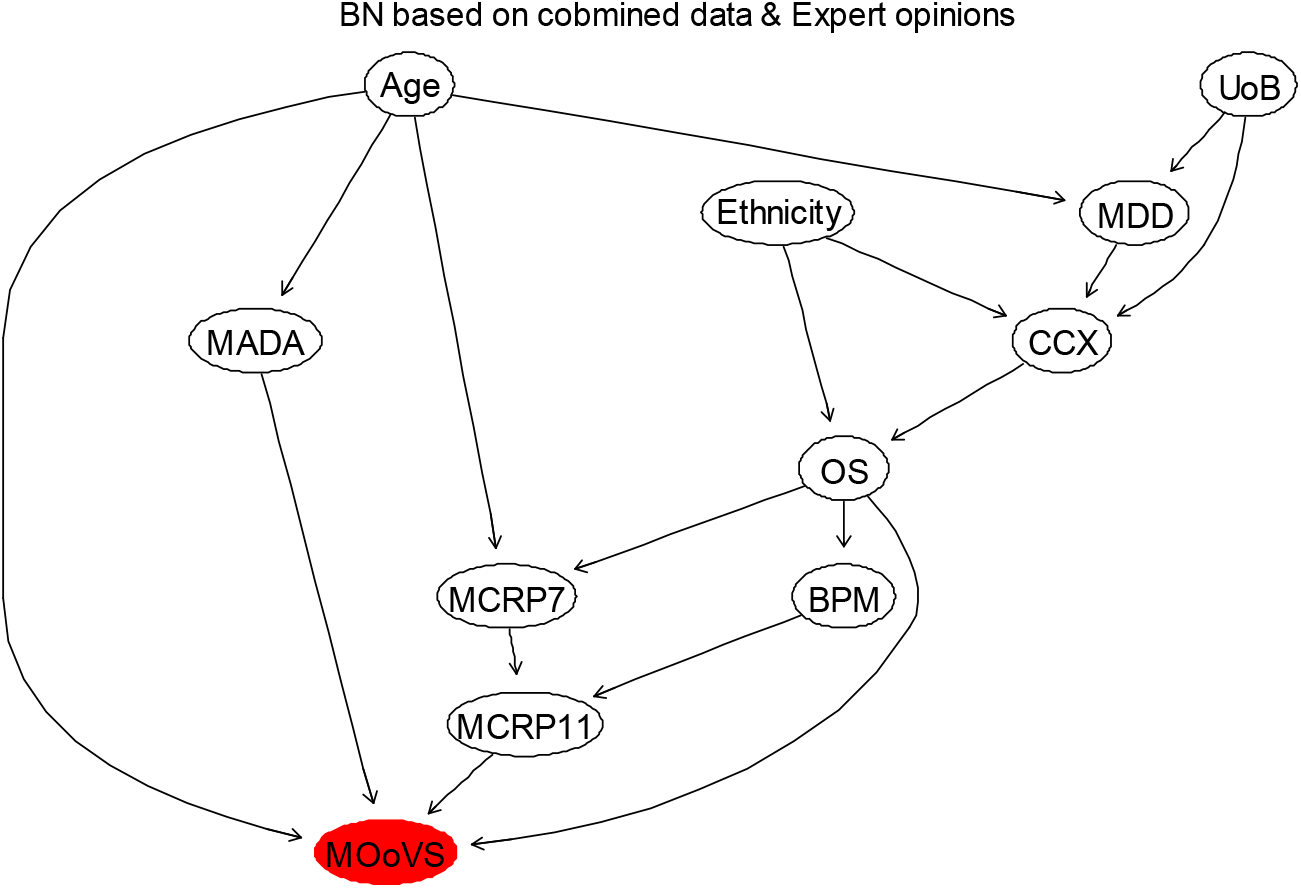
The BN that is learned from the combined data and non-domain expert opinions to model “MOoVS” in terms of the most influencing factors.

**Figure 6.**
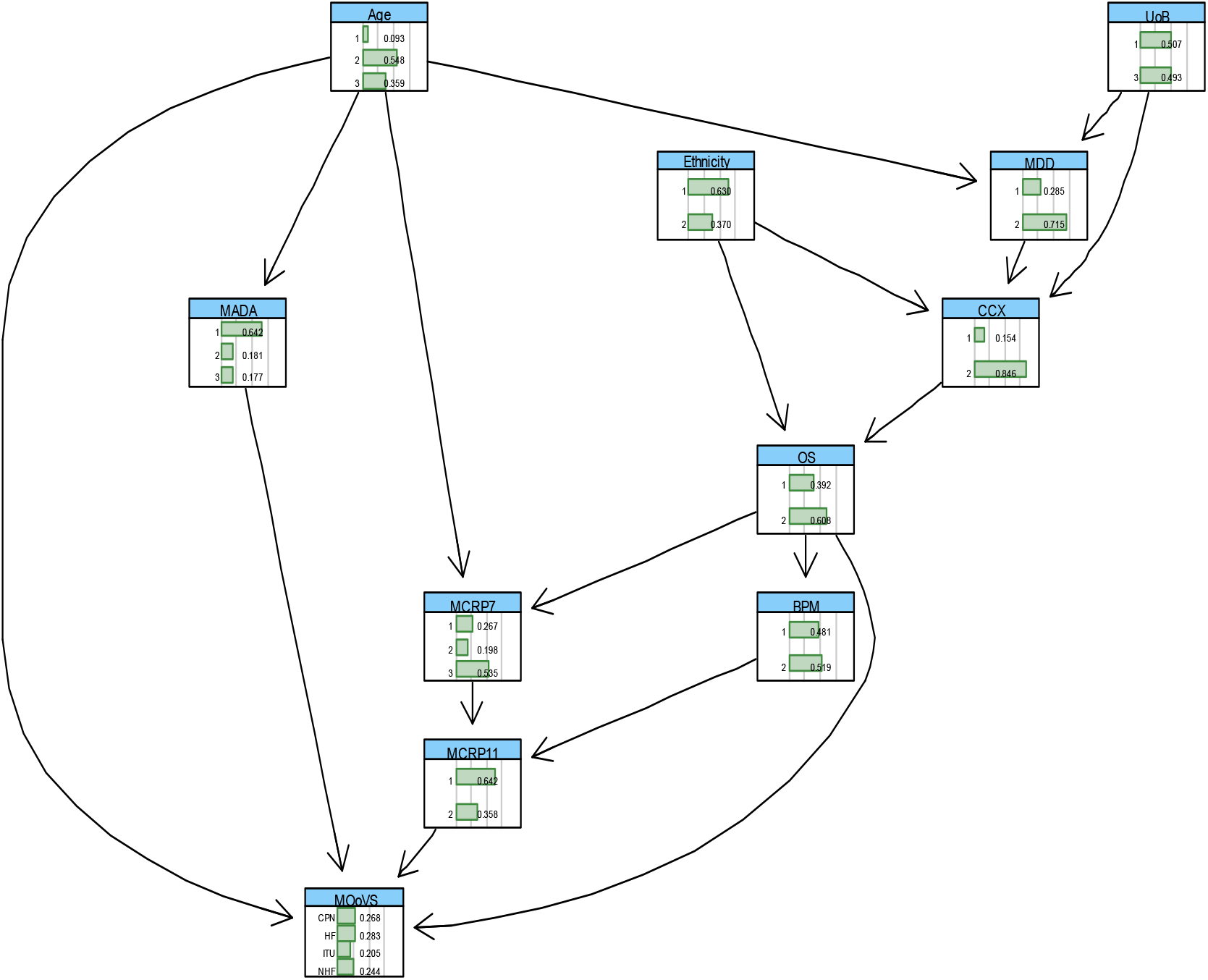
The BN with conditional probability tables learned for “MOoVS” outcome based on the combined elicited domain expert opinions with the (balanced) data.

**Figure 7.**
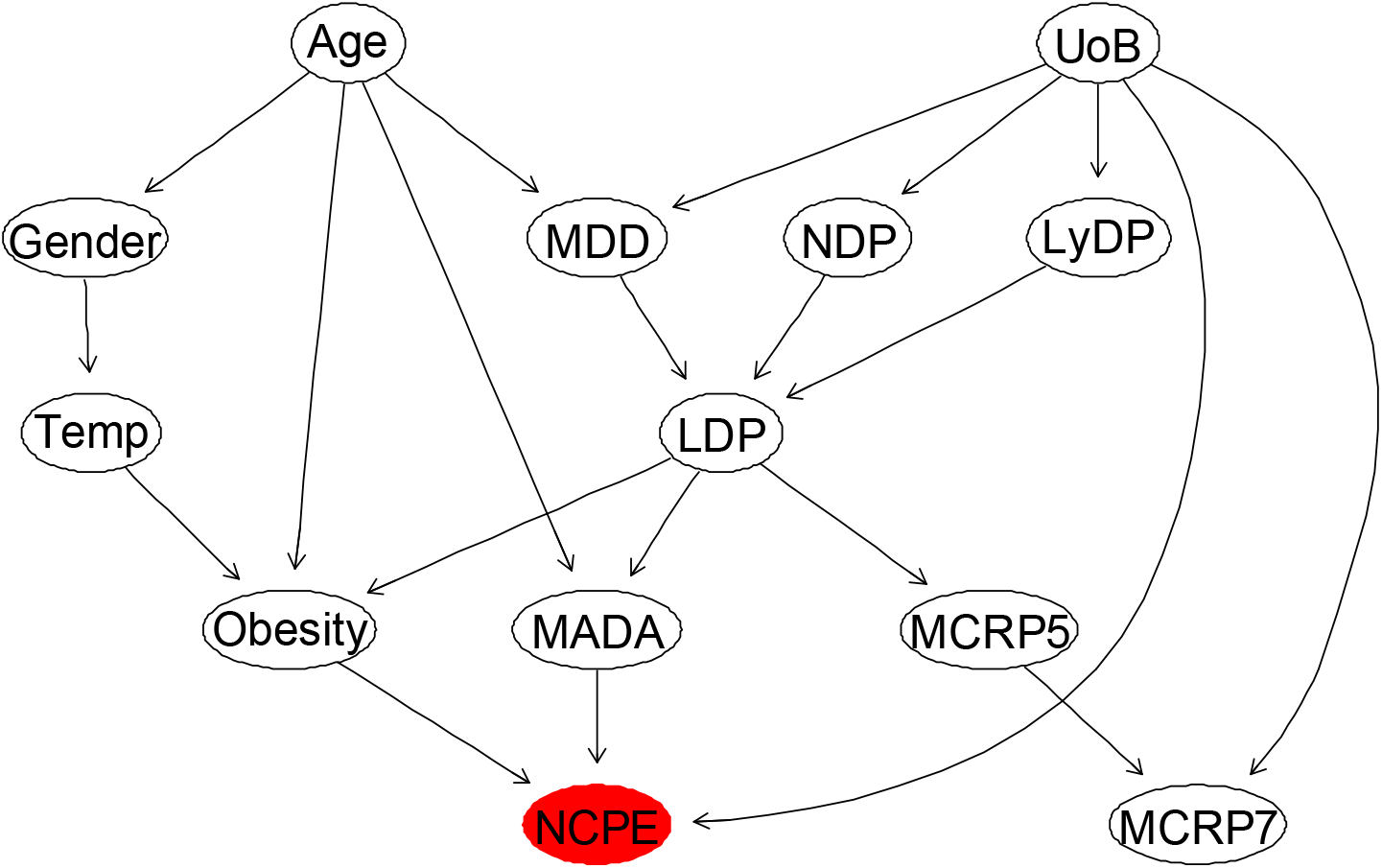
The BN that is learned from the combined data and domain expert opinions to model “NCPE” in terms of its most influencing factors.

**Figure 8.**
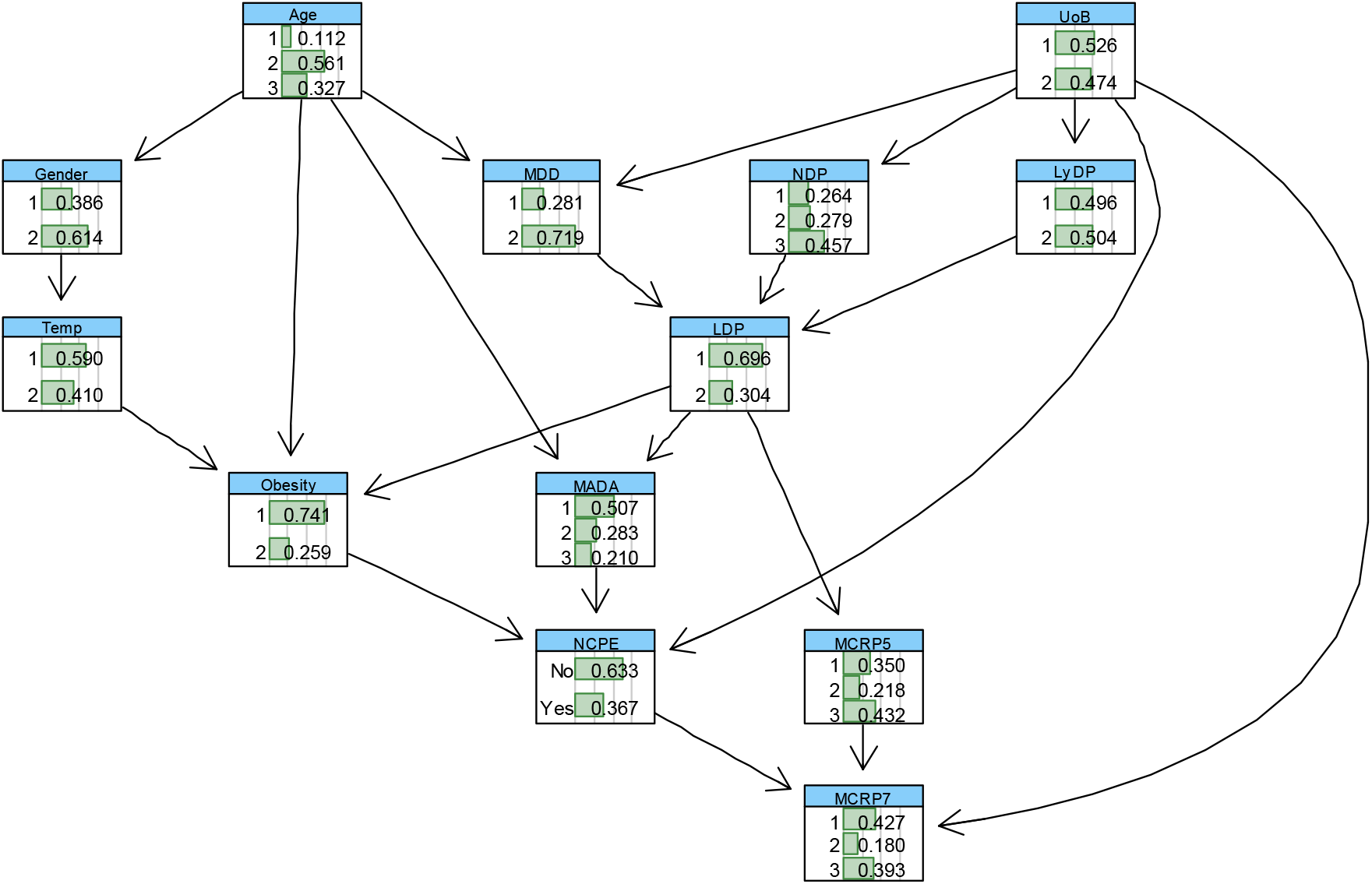
The BN shown in Figure 12, with the computed conditional probability tables learned from data.

Table 10 illustrates several metrics to assess predictive performance of the proposed BN for the 3 different categories of ADT. Examining the raw classification accuracy is the first step in assessing the performance of a model. This can be done by computing the overall classification accuracy rate that corresponds to the proportion of observations that have been correctly classified. Despite the small sample size and high percentages of missing values of the raw data, the computed overall accuracy of 61.5% is very promising. However, the original data transformed to be balanced with respect to the ADT outcome variable, but the ADT variable and the selected test data set are still quite imbalanced. As a result, the balanced accuracy for each state of ADT was computed which are reported in Table 10. The lowest balanced accuracy is computed for ‘2 ≤ *ADT* < 3^’^ days which indicates that this class represented a minority in comparison to the other classes. The computed sensitivity measures, which is the metric to evaluate the learned BN ability to predict true positives of each available category of ADT, suggest that “ADT ≥ 3” days has the highest rate (77%), and ‘2 ≤ *ADT* < 3^’^ days has the lowest sensitivity rate (41.4%). We also compute specificity, which is the metric to evaluate the fitted BN ability to predict true negatives of each ADT category. The results suggest “ADT ≤ 1” days (83.5%) and “ADT ≥ 3” days are the categories with the highest and lowest specificity rates, respectively. The next important metric is F1-score that can be interpreted as a weighted average of the precision and sensitivity values, where an F1 score reaches its best value at 1 and worst value at 0. Since, the F1-score takes both false positives and false negatives into account; it will be usually more useful than accuracy, especially if the original test dataset has an uneven class distribution. The computed F1-scores for the ADT categories suggest promising accuracy for ‘ADT ≤ 1’ days and ‘ADT ≥ 3’ days.

**Table 10.**
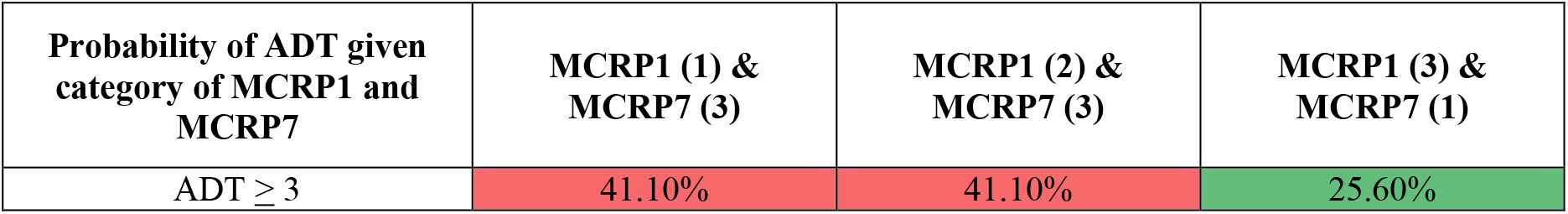
The heat-mapped, conditional probabilities of ADT given MCRP1 and MCRP7 levels. The ADT categories are ‘1’ (≤ <u>1</u> day), ‘2’ (<2 days but < 3 days) and ‘3’ (≥ 3 days). MCRP1 has been divided into 3 categories: ‘1’ (≤ 30), ‘2’ (31-100) and ‘3’ (≥100). MCRP7 has been divided into 3 categories: ‘1’ (≤ 50), ‘2’ (51-100) and ‘3’ (≥100).

Understanding the simultaneous impact of MADA, obesity, and MCRP1 on the duration of COVID-19 treatment in patients is increasingly important to manage the growing, unrelenting pressures on hospitals and the NHS. Table 11 shows the probabilities of several important queries computed from the learned BN illustrated in Figure 8. As it is evident from this table, the duration of treatment of 71% of non-obese COVID-19 patients with normal MADA levels (≥35 g/L) and low MCRP1 (≤30 mg/L) is up to one day. In addition, the treatment duration of 95% of the patients with the above characteristics would be less than 3 days. In comparison to the obese patients, it can be observed this probability (i.e., probability that the duration of treatment is up to one day) will be reduced to 54.4% (with the same characteristics). On the other hand, the predicted probabilities for duration of treatment of the obese and on-obese patients, with very low levels of MADA (≤30 g/L), regardless of levels of MCRP1, are not significantly different from each other. Furthermore, the model and results reported in Table 11 suggest that the levels of MCRP1 alone would not be adequate to accurately predict the probabilities of treatment duration of more than 3 days. These probabilities must be updated by adding more evidence about the levels of MCRP at other days. It would be straightforward to revise and update the BN illustrated in Figure 8 by augmenting the other outcomes, for example “IPD”, to understand what proportion of patients with a treatment duration ≤ 3 days may not survive. Overall, it can be observed that the COVID-19 treatment duration is higher for obese patients with high CRP levels and low Albumin levels.

Similar to the IPD outcome, it is of great interest to understand how the trend in CRP at different days along the course of COVID-19 infection, in addition to the individual CRP levels, can be incorporated and evaluated using the learned BN for ADT, as shown in Figure 8. By a similar argument discussed above, the conditional probabilities of ADT given CRP levels at different days are computed for the ADT variable. Table 12 illustrates the conditional probabilities of ADT given different levels of MCRP1 and MCRP7. The results provided in this table suggest that if there is an increasing gradient from MCRP1 to MCRP7, which represents a worsening infection, the probability that the patient will require at least 3 days of treatment will consequently increase from 28% to 41%. On the contrary, if there is a decreasing gradient from MCRP1 to MCRP7, which represents a resolving infection, the probability that the patients require at least 3 days of treatment will correspondingly decrease from the early prediction of 37%, to less than 26%.

### 3.3 Modelling Max Oxygen or Ventilatory Support (MOoVS) using Bayesian networks

Modelling MOoVS based on the provided data using a BN model was very challenging, as this variable was severely imbalanced; Not requiring High Flow Oxygen (NHF) (63.4%), requiring High Flow Oxygen (HF) (25.9%), requiring CPAP/NIV (CPN) (4.5%) and requiring ITU Admission (ITU) (6.2%). The BN was first fitted to this imbalanced and small dataset, but the predictive performance metrics were poor. In order to resolve this issue, we use the ML-based technique known as “SMOTE” which is briefly explained in Subsection 2.61, to make data balanced with respect to “MOoVS”. The results presented in this section are derived after the MOoVS variable was balanced using the SMOTE method.

We then conduct the feature selection approaches as described in Section 2.62, to identify the most important risk factors affecting MOoVS. The results of the feature selection are reported in Table 3, Figure 1, and Figure 16 in appendix A. The first 10 most relevant independent variables affecting MOoVS were then selected based on the feature selection results (Table 3) to learn a BN for MOoVS. This BN was learned based on the observed processed data, combined with domain expert opinions about the network structure and model predictions. The performance of the resulting BN model was critically examined and compared against other suitable BN candidates which were learned based on the observed data only, using several model diagnostic algorithms, including k-fold cross validation. The BN model illustrated in Figure 10, explaining interdependencies between MOoVS and the most relevant independent variables, is the final selected model after critical model checking and considering the domain experts’ feedbacks.

Table 13 illustrates several metrics to assess predictive performance of the proposed BN for the different states of “MOoVS”. The overall classification accuracy suggests that over 60% of cases have been correctly classified. Despite the small sample size of data and high percentages of missing values of the raw data, the computed overall accuracy (60.25%) is quite promising. As stated above, the original imbalanced database was transformed using the SMOTE method to balance the dataset with respect to the ADT outcome variable. However, the resulting MOoVS variable (comprising of NHF (24.5%), HF (28%), CPN (27%) and ITU, 20.5%), and the selected test dataset are still quite imbalanced. As a result, the balanced accuracy for each category of MOoVS is computed and reported in Table 13. It can be observed that the balanced accuracy of each state is significantly improved (compared to the overall accuracy), varying from 63.6% (“CPN”) to 80.7% (“ITU”). The ability to predict true positives of each available category of MOoVS is measured by Recall, or Sensitivity, which suggests “ITU” has the highest rate (89%), and “CPN” has the lowest sensitivity rate (36%). We also compute specificity whereby “CPN” (91%) and “ITU” (73%) are the categories with the highest and lowest rates, respectively. Since the original raw data has an uneven class distribution, F1-score as a weighted average of the precision and sensitivity values would be very useful. The computed F1-scoures for most of MOoVS categories, except “High flow O_2_” (56%) are promising.

**Table 113.**
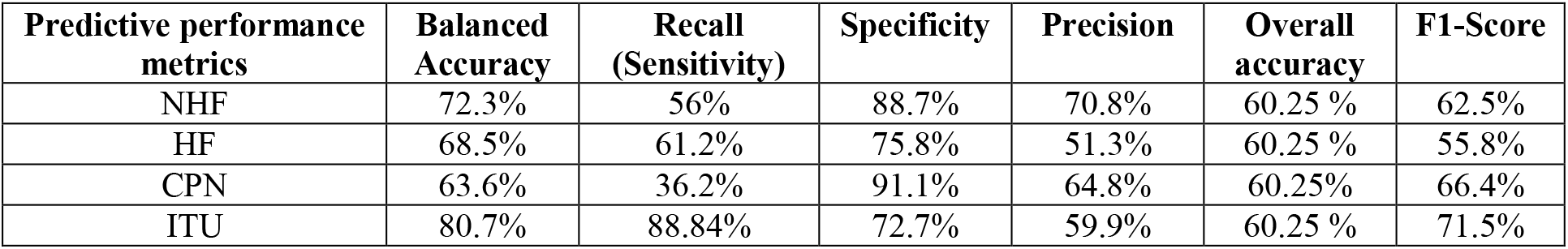
Summary of the predictive performance results of BN developed to model MOoVS as illustrated in Figure 10. The 4 categories of MOoVS were no high-flow oxygen (NHF), high-flow oxygen (HF), CPAP/NIV (CPN) or ITU admission (ITU).

It is of great important to understand how the right level of Max oxygen (HF, or NHF), or ventilatory support should be selected, depending on the level of MADA, state of Oxygen Saturations (OS), and levels of MCRP11, to enhance the survival rate and recovery speed of COVID-19 patients. Table 14 shows the probabilities of requiring different levels of oxygen or ventilatory support given different states of MADA, OS and MCRP11 levels. The illustrated results in this table suggest that most of the patients with the better health characteristics such as high MADA (≥35 g/L), high OS (≥ 92%), and low MCRP11 (≤100 mg/L) are likely to only require no-high flow O_2_ (73%) or high flow O_2_ (12.5%). These patients are thus suitable for ward-based care and will not need ITU admission. On the contrary, when MADA and OS levels respectively drop to below 30 g/L and 91%, and MCRP11 level increases to over 100 mg/L, the need is increased for these patients to receive CPAP/NIV (37%) or ITU admission (49%).

**Table 14.** The heat-mapped, conditional probabilities of MOoVS given the different configurations of MADA (1= ‘≤30’, 2= ‘30-35’, 3= ‘35≤‘), OS (‘1’ = ≥92 and ‘2’ <92) and MCRP11 (1= ≤100 and 2= >100). Patients either required no high-flow oxygen (NHF), high-flow oxygen (HF), CPAP/NIV (CPN) or ITU admission (ITU).

| Probability of MOoVS given category of OS, MADA & MCRP11 | OS (1), MADA (3) & MCRP11 (1) | OS (1), MADA (1) & MCRP11 (2) | OS (1), MADA (1) & MCRP11 (1) | OS (2), MADA (3) & MCRP11 (1) | OS (2), MADA (1) & MCRP11 (1) | OS (2), MADA (1) & MCRP11 (2) |
| --- | --- | --- | --- | --- | --- | --- |
| NHF | 72.80% | 39.30% | 34.80% | 20.10% | 9.80% | 1.80% |
| HF | 12.50% | 10.10% | 38.90% | 45.90% | 25.40% | 12.40% |
| CPN | 14.70% | 26.90% | 18.60% | 34% | 39.30% | 36.90% |
| ITU | 0% | 23.70% | 7.70% | 0% | 25.50% | 48.90% |

The probabilities given in Table 14 can be significantly altered in the light of new evidence, such as patient age. The updated probabilities are shown below in Table 15. For example, all young patients with OS ≥92, MADA≤30 & MCRP11≤100, require either NHF (80%) or HF (20%) to recover. If MADA levels of young patients increase to over 35 g/L, with the same levels of OS (≥92) and MCRP11 (≤ 30), they will more likely need to use only NHF (93%) to recover. However, if the health of the patients start to deteriorate, as MADA and OS levels respectively drop to ≤30 g/L and <92%; and MCRP11 level increases to over 100 mg/L, their need for HF, CPN or ITU will significantly increase depending to the age of patient. For the young patients, CPAP/NIV (36.5%) or ITU (63.5%) would be recommended. However, for patients ≥ 70 years, either HF (22%) or CPAP/NIV (76%), and paradoxically not ITU (0%), would be recommended usually because they are deemed unsuitable for ITU admission due to the futility of ITU-based treatment relative to a younger patient. This reflects the rationing of healthcare resources that occurs in hospital after difficult medico-ethical decisions.

**Table 15.**
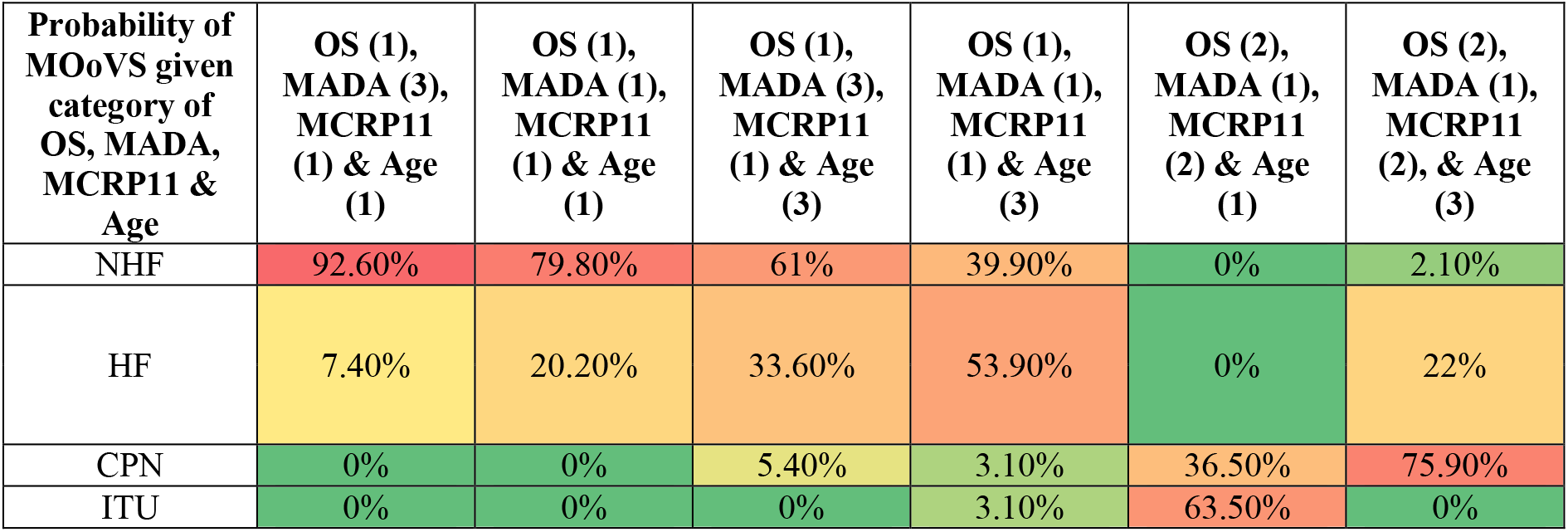
The heat-mapped, conditional probabilities of MOoVS given the different configurations of Age (1= ‘≤40 years’ and 2= ‘≥70 years’) MADA (1= ‘≤30’, 2= ‘30-35’, 3= ‘35≤‘), OS (‘1’ = ≥92 and ‘2’ <92) and MCRP11 (1= ≤100 and 2= >100). Patients either required no high-flow oxygen (NHF), high-flow oxygen (HF), CPAP/NIV (CPN) or ITU admission (ITU).

| Probability of MOoVS given category of OS, MADA, MCRP11 & Age | OS (1), MADA (3), MCRP11 (1) & Age (1) | OS (1), MADA (1), MCRP11 (1) & Age (1) | OS (1), MADA (3), MCRP11 (1) & Age (3) | OS (1), MADA (1), MCRP11 (1) & Age (3) | OS (2), MADA (1), MCRP11 (2) & Age (1) | OS (2), MADA (1), MCRP11 (2), & Age (3) |
| --- | --- | --- | --- | --- | --- | --- |
| NHF | 92.60% | 79.80% | 61% | 39.90% | 0% | 2.10% |
| HF | 7.40% | 20.20% | 33.60% | 53.90% | 0% | 22% |
| CPN | 0% | 0% | 5.40% | 3.10% | 36.50% | 75.90% |
| ITU | 0% | 0% | 0% | 3.10% | 63.50% | 0% |

### 3.4 Modelling New Confirmed Pulmonary embolism during Admission (NCPE) using Bayesian networks

Modelling the new confirmed pulmonary embolism during admission (NCPE) based on the massively incomplete and imbalanced data using BN models, similar to modelling of other outcomes of interests discussed above, was very challenging. In order to resolve this issue, we used the ML-based technique known as “SMOTE” which is briefly explained in Subsection 2.61, to balance data with respect to “NCPE”. We then perform the feature selection approaches as described in Section 2.62, to identify the most important risk factors affecting the NCPE. The results of the feature selection are reported in Table 3, Figure 1, and Figure 17 in appendix A. The first 12 most relevant independent variables affecting NCPE were then selected based on the reported feature selection results, to learn a BN model for NCPE. This BN was learned based on the observed processed and balanced data, combined with the expert opinions elicited from the domain experts about the network structure and the effective covariates on the NCPE. The performance of the resulting BN model was critically examined and compared against other suitable BN candidates (which were learned by including the observed data only), using several model diagnostic algorithms, including k-fold cross validation. The BN model illustrated in Figure 12, explaining interdependencies between NCPE and the most relevant independent variables, is the final selected model after critical model checking and considering the domain experts’ feedbacks.

The augmented BN shown in Figure 12, with added computed conditional probability tables learned from data, is illustrated in Figure 13. The marginal probability table of each variable, after the whole dataset was balanced with respect to the NCPE, can be observed on each node. We critically explore the predictive behaviour of the NCPE with respect to various simultaneous changes of the independent variables, selected according to the hypothesis of interest.

We first use different metrics, including PPV, NPV, sensitivity, specificity, overall accuracy, and F-Score, to evaluate the predictive performance of the BN suggested to model the NCPE in terms of the related factors as illustrated in Figure 12. The values of these metric for the learned BN for NCPE are computed in Table 16.

**Table 16.** Summary of predictive performance results of the BN learned for the NCPE as shown in Figure 13.

| Predictive performance metric | PPV | NPV | Specificity | Sensitivity | Overall accuracy | F1-Score |
| --- | --- | --- | --- | --- | --- | --- |
| BN for IPD | 83.7% | 80.9% | 75% | 87.9% | 82.7% | 85.8% |

The computed F1-score of almost 86% shows the classification prediction of the learned BN for NCPE is quite precise and is robust (in the sense that it does not miss a significant number of instances). In addition, the PPV (83.7%), sensitivity (88%) and NPV (80.9%), which collectively represent the BN model’s ability to predict inpatient NCPE, was high.

One of the hypotheses of interest is whether there is a significant association between the extent of COVID-19 changes on CT scan (UoB) with NCPE. Table 17 shows the conditional probability of NCPE given the different states of UoB. The reported results in this table suggest that there is strong association between “bilateral” states of UoB with NCPE, based on the computed conditional probability, of 72.4%. This suggests that more extensive ground-glass or consolidative changes on CT scan secondary to COVID-19 may be associated with the development of PE sequelae secondary to COVID-19.

**Table 17.**
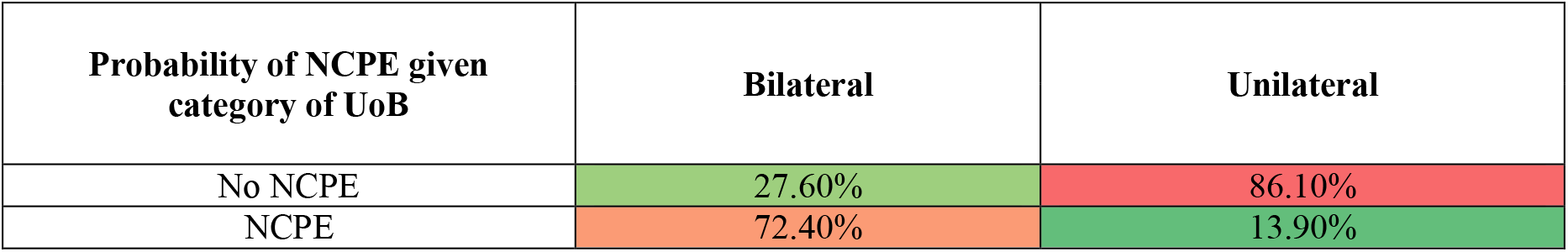
The heat-mapped, conditional probabilities of NCPE given the different states of UoB (either unilateral or bilateral CT scan changes).

| Probability of NCPE given category of UoB | Bilateral | Unilateral |
| --- | --- | --- |
| No NCPE | 27.60% | 86.10% |
| NCPE | 72.40% | 13.90% |

Another research question that can be verified using the BN developed for the NCPE is to explore whether there is a significant association between the maximum D-Dimer during admission (MDD levels) with NCPE. Table 18 shows the conditional probability of NCPE given the different states of MDD. It can be concluded that there is a moderate association between high values of MDD with NCPE (54.5%).

**Table 18.** The heat-mapped, conditional probabilities of NCPE given the different states of MDD.

| Probability of NCPE given category of MDD | MDD |  |
| --- | --- | --- |
| | $\leq 400$ | $400 <$ |
| No NCPE | 65.40% | 45.50% |
| NCPE | 34.60% | 54.50% |

Table 19 reports the joint impact of UoB and MDD on NCPE. The results suggest that the presence of NCPE is more significantly influenced by the presence of bilateral ground-glass or consolidative CT scan changes secondary to COVID-19, rather than levels of MDD. This may be explained by the fact that many other conditions can also increase D-Dimer such as disseminated intravascular coagulation, deep vein thrombosis and infection, which could thus be leading to high rates of PE false positives amongst COVID-19 patients. Furthermore, not all patients with raised D-dimers would have had CT-scans to investigate for PE, especially if it was deemed futile and the patient was palliative.

**Table 19.** The heat-mapped, conditional probabilities of NCPE given the different states of MDD and UoB.

| Probability of NCPE given category of UoB & MDD | Bilateral CT changes & MDD $\leq 400$ | Bilateral CT changes & MDD $> 400$ | Unilateral CT changes & MDD $\leq 400$ | Unilateral CT changes & MDD $> 400$ |
| --- | --- | --- | --- | --- |
| No NCPE | 27.20% | 27.80% | 86.50% | 85.90% |
| NCPE | 72.80% | 72.20% | 13.50% | 14.10% |

Table 20 highlights the combined impact of various configurations of UoB and MADA levels on the NCPE. The results suggest that the presence of NCPE is significantly influenced by bilateral CT involvement, and MADA ≤30.

**Table 20.** The heat-mapped, conditional probabilities of NCPE given the different states of MADA and UoB.

| Probability of NCPE given categories of UoB & MADA | Bilateral CT changes & MADA $\leq 30$ | Bilateral CT changes & MADA $\geq 35$ | Unilateral CT changes & MADA $\leq 30$ | Unilateral CT changes & MADA $\geq 35$ |
| --- | --- | --- | --- | --- |
| No NCPE | 27% | 51% | 85.30% | 98.60% |
| NCP | 73% | 49% | 14.70% | 1.40% |

We finally investigate the impact of obesity alongside of MADA and MCRP7 levels on NCPE. Table 21 illustrates the conditional probabilities of NCPE given various configurations of interest of MADA and MCRP7 levels for obese and non-obese patients. It can be concluded that there is a strong association (70%) between the presence of NCPE and MADA ≤30, as well as MCRP7 ≥100, for the non-obese patients. If the patient is obese, MADA ≤30 seems to be more influential in contributing towards the risk of NCPE, as opposed to MCRP7 ≥ 100.

**Table 21.** The heat-mapped, conditional probabilities of NCPE given the different states of MADA, MCRP7 and Obesity.

| Probability of NCPE given categories of MADA, MCRP7 and Obesity | MADA (1) & MCRP7 (1) | MADA (1) & MCRP7 (3) | MADA (3) & MCRP7 (1) | MADA (3) & MCRP7 (3) |
| --- | --- | --- | --- | --- |
| **BMI < 30 (Non-Obese Patients)** |  |  |  |  |
| No NCPE | 44.20% | 30.90% | 63% | 44.70% |
| NCPE | 55.80% | 69.10% | 37% | 55.30% |
| **BMI $\geq 30$ (Obese Patients)** |  |  |  |  |
| No NCPE | 68.20% | 51.50% | 94.30% | 87.80% |
| NCPE | 31.80% | 48.50% | 5.70% | 12.20% |

## 4. Discussion

In this study, in addition to quantifying the significance of feature-selected risk factors, we showcase the use of Bayesian Networks to accurately predict four different COVID-19 inpatient outcomes: inpatient mortality (IPD), maximum level of oxygen or ventilatory requirement (MOoVS), duration of inpatient COVID-19 treatment (ADT), and new confirmed diagnoses of pulmonary embolism (NCPE), all whilst using a relatively small sample-size. The models can predict these outcomes using different combinations of readily available clinical data which serve as the independent predictor variables, whilst also accounting for interdependency between these variables.

Various COVID-19 prognostic indicators have been described in the literature, such as neutrophil: lymphocyte ratio, C-Reactive Protein (CRP), age, gender, ethnicity, oxygen saturation on admission, diabetes mellitus, hypertension, malignancy, obesity and COPD [39]. However, drawing insights from this information is impeded by the lack of clarity as to the relative influence each of these indicators has on mortality. In the clinical setting, patients often present with different combinations of these risk factors and biomarkers. Consequently, weighing them all up in order to allocate scarce healthcare resources can be challenging. This highlights a role for a predictive, quantitative risk-stratification tool. Our model has been constructed so that it can utilize data at first clinical presentation, for example, in the emergency department, but also after 3 days, and 7 days of inpatient treatment. This can allow clinicians to risk stratify at different time-points during an inpatient stay. In addition, our model can also predict the duration of inpatient COVID-19 treatment and maximum level of oxygen requirement that a patient may need during their inpatient stay. This may aid emergency physicians with the decision as to whether to admit a patient to hospital and avoid failed discharges, but also medical physicians with the decision as to whether to refer to ICU prior to a patient’s clinical deterioration. The predicted duration of inpatient COVID-19 treatment can be especially useful for bed managers in orchestrating patient flow which is essential to prevent the growing problem of hospital acquired COVID-19 secondary cross-contamination [40]. Furthermore, within the context of hospitals which have reached their maximum ITU capacity, by using our predictive model to identify high-risk patients earlier on in their disease course, clinicians can transfer these high-risk patients to neighbouring hospitals prior to their clinical deterioration.

One of the major strengths of our machine learning based predictive model is that it can predict 4 outcomes. This increases the amount of clinical utility that can be offered to guide clinical decision making. Secondly, this data set has incorporated 44 different variables from 355 patients who all received the primary study outcomes, reducing the risk of confounding error and selection bias. Another strength of this study is that data extraction was conducted and checked manually by trained medical physicians rather than by using coding. It has been well documented that hospital clinical coding is still not entirely accurate within the UK [41,42] and therefore insights drawn from national databases may be prone to significant information bias and thus systematic error. Furthermore, it has now been estimated that the sensitivity of the SARS CoV-2 RT-PCR nasopharyngeal swab is likely to be 50-75% [43–47]. This has created a huge global problem in diagnosing and identifying COVID-19 patients, especially because not all patients exhibit symptoms [48–52]. To overcome this issue, the cohort of patients with COVID-19 who had negative RT-PCR swabs but positive CT scan imaging (n=55) were also included in this study.

The biggest limitation to our study is that since the study was conducted in a single centre, not only is the n number limited, but the data only reflects the demographics of the surrounding population. Within the UK, geographical location and socio-economic factors are heavily influencing death rates [53] and, thus, our data may have limited generalizability to the wider UK population by not accounting for these factors. Furthermore, our data has only been collected from hospital inpatients and therefore our model is not generalizable to the wider community either. Secondly, data was not always available, or accurate, for all patients. This was sometimes due to a lack of documentation, usually if the attending physicians at the time did not deem the information relevant, or if the information was not available, especially in patients who were cognitively impaired without any next of kin to provide collateral histories. Moreover, not all investigations, such as CT scans, were required for every patient and may have not been done due to the limited resources available in the NHS. Subsequently, only patients deemed to have abnormal results would have been the patients to receive the investigation. Also, with regards to palliation, some patients clearly had different treatment goals to others. For example, this would suggest that patients with severe COVID-19 who could have had ventilatory support may have not had it because their treatment goal was palliation instead. All these factors together introduce information bias secondary to data measurement. Finally, although most parameters were objectively documented on the electronic patient record system, some data, such as ethnicity, was self-reported by patients thus also introducing a modest element of recall bias.

## 5. Conclusion

Overall, our findings demonstrate reliable, multivariable predictive models for 4 outcomes, that utilize readily available clinical information for COVID-19 adult inpatients. The models, if provided with more training data, have the potential to be refined even further. Future research is required to externally validate our models and demonstrate their utility as clinical decision-making tools.

## Data Availability

Anonymized data regarding 44 independent predictor variables of 355 adults diagnosed with COVID-19, at a UK hospital, was manually extracted from electronic patient records for retrospective, case-controlled analysis.

## 6. Acknowledgements

None

## Abbreviations

(OS): Oxygen Saturations
(BPM): Respiratory Rate
(UoB): CT imaging severity of COVID-19 related changes
(CCX): COVID-19 related Chest X-Ray changes
(MADA): Albumin
(MDD): D-Dimer
(CRP): C-Reactive Protein
(MCRP1): CRP Day 1-2
(MCRP3): CRP Day 3-4
(MCRP5): CRP Day 5-6
(MCRP7): CRP Day 7-8
(MCRP11): CRP Day 11-12
(IPD): Inpatient Mortality
(MOoVS): Maximum Oxygen or Ventilatory Support
(ADT): Duration of Treatment for COVID-19
(NCPE): New confirmed diagnosis of pulmonary embolism

## Appendix A

**Table A.1:** Patient Characteristics and descriptive statistics

| Demographics (n=355) |  |  |  |  |  |  |
| --- | --- | --- | --- | --- | --- | --- |
| Male (%) | 52 |  |  |  |  |  |
| Non-Caucasian (%) | 30 |  |  |  |  |  |
| Age (Years) | Median | Upper Quartile | Lower Quartile | Interquartile Range | Mean | Standard Deviation |
|  | 65 | 81 | 52 | 29 | 65 | 18 |
| Vital Observations (n=355) |  |  |  |  |  |  |
|  | Median | Upper Quartile | Lower Quartile | Interquartile Range | Mean | Standard Deviation |
| Oxygen Saturation (%) | 93 | 96 | 89 | 7 | 91 | 8 |
| Respiratory Rate (breaths per minute) | 24 | 28 | 20 | 8 | 25 | 7 |
| Temperature (°Celsius) | 37.9 | 38.6 | 37.1 | 1.5 | 37.9 | 1.0 |
| Comorbidities and Medications (%) (n=355) |  |  |  |  |  |  |
| Obesity |  |  | 34.3 |  |  |  |
| Previous Venous Thromboembolism |  |  | 6.5 |  |  |  |
| Chronic Obstructive Pulmonary Disease |  |  | 11.5 |  |  |  |
| Bronchiectasis |  |  | 2 |  |  |  |
| Asthma |  |  | 9.9 |  |  |  |
| Interstitial Lung Disease |  |  | 2.3 |  |  |  |
| Lung Cancer |  |  | 2.5 |  |  |  |
| Neuromuscular Disease |  |  | 15.8 |  |  |  |
| Diabetes Mellitus |  |  | 24.2 |  |  |  |
| Hypertension |  |  | 36 |  |  |  |
| Ischaemic Heart Disease |  |  | 12.1 |  |  |  |
| Chronic Kidney Disease |  |  | 13.2 |  |  |  |
| Non-steroidal anti-inflammatory drugs |  |  | 3.1 |  |  |  |
| Anticoagulant |  |  | 16.9 |  |  |  |
| Antibiotics |  |  | 3.1 |  |  |  |
| Immunosuppressants |  |  | 5 |  |  |  |
| Corticosteroids |  |  | 4.2 |  |  |  |
| Angiotensin Converting Enzyme Inhibitors |  |  | 15.2 |  |  |  |
| Angiotensin Receptor Blockers |  |  | 6.8 |  |  |  |
| Radiographic Changes (%) |  |  |  |  |  |  |
| Bilateral COVID-19 CT changes (n=117) |  |  | 91.5 |  |  |  |
| COVID-19 Chest X-Ray Changes (n=341) |  |  | 69.5 |  |  |  |
| Biochemical Markers |  |  |  |  |  |  |
|  | Mean | Median | Upper Quartile | Lower Quartile | Interquartile Range | Standard Deviation |
| Lactate (mmol/L), n=242 | 1.85 | 1.4 | 2 | 1.1 | 0.9 | 1.63 |
| Lymphocytes (10<sup>9</sup>/L), n=351 | 1.10 | 1 | 1.3 | 0.7 | 0.6 | 0.67 |
| Neutrophils (10<sup>9</sup>/L), n=351 | 6.75 | 5.4 | 8.8 | 3.25 | 5.55 | 4.86 |
| Albumin (g/L), n=341 | 31.38 | 31 | 36 | 27 | 9 | 6.6 |
| Ferritin (ng/L), n=51 | 866.33 | 725 | 1371.5 | 264.5 | 1107 | 760.43 |
| D-Dimer (ng/ml), n=115 | 2604.11 | 656 | 2163 | 344.5 | 1818.5 | 7016.26 |
| Fibrinogen (g/L), n=18 | 44.91 | 5.85 | 8.56 | 4.41 | 4.15 | 121.97 |
| Troponin (Local units), n=75 | 628.2 | 30.9 | 90.9 | 7.35 | 83.55 | 3201.81 |
| C-Reactive Protein (CRP) Day 0 (mg/L), n=337 | 110.39 | 96 | 164 | 39 | 125 | 89.13 |
| CRP Day 1-2 (mg/L), n=217 | 152.36 | 142 | 212 | 77 | 135 | 94.84 |
| CRP Day 3-4 (mg/L), n=185 | 146 | 117 | 203 | 67 | 136 | 106.14 |
| CRP Day 5-6 (mg/L), n=139 | 132.6 | 111 | 187 | 55.5 | 131.5 | 103.54 |
| CRP Day 7-8 (mg/L), n=106 | 117.92 | 87.5 | 164 | 46 | 118 | 99.67 |
| CRP Day 9-10 (mg/L), n=75 | 122.65 | 98 | 170.5 | 51.5 | 119 | 95.25 |
| CRP Day 11-12 (mg/L), n=63 | 94.46 | 73 | 124.5 | 33.5 | 91 | 83.15 |
| CRP Day 13-14 (mg/L), n=43 | 91.6 | 65 | 121 | 38.5 | 82.5 | 77.49 |
| CRP Day 15-20 (mg/L), n=36 | 97.43 | 57.5 | 133.5 | 26.5 | 107 | 91.67 |

